# Low-dose interleukin 2 antidepressant potentiation in unipolar and bipolar depression: Safety, efficacy, and immunological biomarkers

**DOI:** 10.1101/2023.09.12.23295407

**Authors:** Sara Poletti, Raffaella Zanardi, Alessandra Mandelli, Veronica Aggio, Annamaria Finardi, Cristina Lorenzi, Giovanna Borsellino, Matteo Carminati, Elena Manfredi, Enrico Tomasi, Sara Spadini, Cristina Colombo, Hemmo A. Drexhage, Roberto Furlan, Francesco Benedetti

## Abstract

Immune-inflammatory mechanisms are promising targets for antidepressant pharmacology. Based on reported immune cell abnormalities, we defined an antidepressant potentiation treatment with add-on low-dose interleukin 2 (IL-2), a T-cell growth factor of proven anti-inflammatory efficacy in autoimmune conditions, increasing thymic production of naïve CD4+ T cells, and possibly correcting the partial T cell defect observed in mood disorders. We performed a single-center, randomised, double-blind, placebo-controlled phase II trial evaluating the safety, clinical efficacy and biological responses of low-dose IL-2 in depressed patients with MDD or BD. 36 consecutively recruited inpatients at the Mood Disorder Unit were randomised in a 2:1 ratio to receive either aldesleukin (12 MDD and 12 BD) or placebo (6 MDD and 6 BD). Active treatment significantly potentiated antidepressant response to ongoing SSRI/SNRI treatment in both diagnostic groups, and expanded the population of Treg, Th2, and Naive CD4+/CD8+ immune cell counts. Changes in cell counts were rapidly induced in the first five days of treatment, and predicted the later improvement of depression severity. No serious adverse effect was observed. This is the first RCT evidence supporting the hypothesis that treatment to strengthen the T cell system could be a successful way to correct the immuno-inflammatory abnormalities associated with mood disorders, and potentiate antidepressant response.

**Highlights:** Immune-inflammatory mechanisms are promising targets for antidepressant pharmacology. In a randomized controlled trial low-dose IL-2 significantly improved antidepressant response. IL-2 rapidly expanded the population of Treg, Th2, and Naive CD4+/CD8+ immune cell counts. Strengthening in the T cell system predicted antidepressant response.

## 1. Introduction

Despite tremendous improvement in antidepressant psychopharmacology based on drugs directly affecting neurotransmitter function, current consensus is that one-third of patients with Major Depressive Disorder (MDD) do not achieve full symptomatic remission, and that in individuals with ineffective initial treatment, many relapses are observed despite continuing apparently effective subsequent treatment, paving the way to treatment-resistant depression (TRD) (Sforzini et al., 2022). Outcomes are even worse in Bipolar Disorder (BD), which has been associated with extremely low success rates of antidepressant drugs in naturalistic settings (Post et al., 2011; Post et al., 2012). Possibly as a consequence of their disabling condition, about 30% of patients with mood disorders attempt suicide (Chen and Dilsaver, 1996; Leverich et al., 2003), and about 20% eventually die from suicide (Osby et al., 2001): hence the need of continuous research on pathogenetic mechanisms to address the clinical need of more precisely targeted and effective antidepressant treatment.

Consistent evidence support immune cellular and inflammatory mechanisms as possible targets for antidepressant pharmacology. *Post-mortem* neuropathology detected increased microglia density and activation, and lymphocyte infiltration, in the brain of suicides (Naggan et al., 2023; Schlaaff et al., 2020; Steiner et al., 2013). Patients with depression show high levels of inflammatory compounds in circulating blood and in CSF; higher baseline immuno-inflammatory setpoints related to circulating cytokines, chemokines, and leukocyte gene expression hamper response to antidepressants; effective antidepressant treatment decreases inflammatory markers; and add-on treatment with immune-modulatory and anti-inflammatory drugs (e.g., minocycline, celecoxib, infliximab) can promote response in TRD (Benedetti and Vai, 2023; Benedetti et al., 2022; Branchi et al., 2021; Kappelmann et al., 2018).

Specific immunopsychiatric antidepressant treatment is however in its infancy, with yet uncertain targets and predictors of response, elusive mechanisms, and a paucity of randomized controlled trials (RCT). Patients with mood disorders show signs of systemic low-grade inflammation due to decreased adaptive, increased innate immunity, with higher macrophage/monocyte inflammatory activation, and higher neutrophils to lymphocyte counts. Immune cellular abnormalities have been described, with a lifetime dynamic pattern of premature immunosenescence and partial T cell defect associated with premature T cell aging. This involves a reduction of naïve CD4+ T cells and an expansion of memory and senescent T cells (Bauer et al., 2015; Becking et al., 2018; Poletti et al., 2017; Simon et al., 2023; Simon et al., 2021a; Swallow et al., 2013). The immune aberrancies trigger a cascade of events which leads to decreased monoaminergic neurotrasmitter function, altered brain glutamatergic activity, and brain structural and functional abnormalities (Benedetti et al., 2020; Bravi et al., 2022; Comai et al., 2022; Haroon et al., 2017; Poletti et al., 2020).

Based on this evidence, we hypothesized that treatments able to strengthen the T cell system could be a successful way to correct these abnormalities, and possibly potentiate antidepressant response. We considered interleukin 2 (IL-2) a good candidate for this purpose. IL-2 is a T-cell growth factor, increasing thymic production of naïve CD4+ T cells even in severe immunodeficiency (Carcelain et al., 2003), and may therefore correct the partial T cell defect observed in mood disorders. IL-2 is essential for CD4+ Th2 differentiation (Cote-Sierra et al., 2004), controls Th1 and Th2 fate decisions in antigen receptor–activated CD4+ T cells (Ross and Cantrell, 2018), and could thus correct the Th1/Th2 shift reported in chronic patients with BD (Brambilla et al., 2014). Starting from 2011 (Koreth et al., 2011; Saadoun et al., 2011), several clinical trials in patients with autoimmune conditions showed that low-dose IL-2 specifically expands and activates CD4+ Treg cell populations and can control inflammation, with a favorable safety profile (Graßhoff et al., 2021; Klatzmann and Abbas, 2015; Rosenzwajg et al., 2019): this effect might also be useful to correct the reported Treg insufficiency in MDD (Ellul et al., 2018; Grosse et al., 2016), thanks to the Treg constitutive expression of high levels of the heterotrimeric high affinity IL-2 receptor complex, which in other CD4+ T cells, CD8+ T cells or NK cells is expressed only upon robust activation (Graßhoff et al., 2021). Finally, IL-2 acts directly as a trophic factor on both neurons and oligodendrocytes (de Araujo et al., 2009), and might then antagonize the detrimental link between low-grade inflammation and brain homeostasis observed in mood disorders.

Low-dose IL-2 was never tested in psychiatric conditions, but in mice it attenuated depression-like behaviors in a chronic stress-induced model of depression (Huang et al., 2022), correcting the Th17/T reg balance and peripheral signs of low grade inflammation. We then performed a randomized, placebo-controlled trial to test low-dose IL-2 as an adjunctive antidepressant treatment in depressed patients with MDD or BD, focusing on alterations in the CD4+ and CD8+ naïve and memory cells, T reg cells, Th17 cells, Th1 cells and Th2 cells, and serum parameters of low grade inflammation (CRP, IL-6), T cell regulation (sCD25, IL-7) and neurotrophism (BDNF).

## 2. Methods

### 2.1 Study design, participants and treatment

The research program on the effects of low dose IL-2 in depression was defined in 2017 in the context of the H2020 MoodStratification project (Drexhage, 2018), and articulated in two parallel trials, one to be conducted in MDD and in BD at the Mood Disorder Unit of Ospedale San Raffaele with the commercially available Aldesleukin (Acronym: IL-2REG; EudraCT N.: 2019-001696-36), and the other in BD at the Assistance Publique - Hôpitaux de Paris with ILT101 (Acronym: DEPIL-2, ClinicalTrials.gov: NCT04133233). Here we present the outcomes of the IL-2REG trial (**Figure 1**).

**Figure 1.**
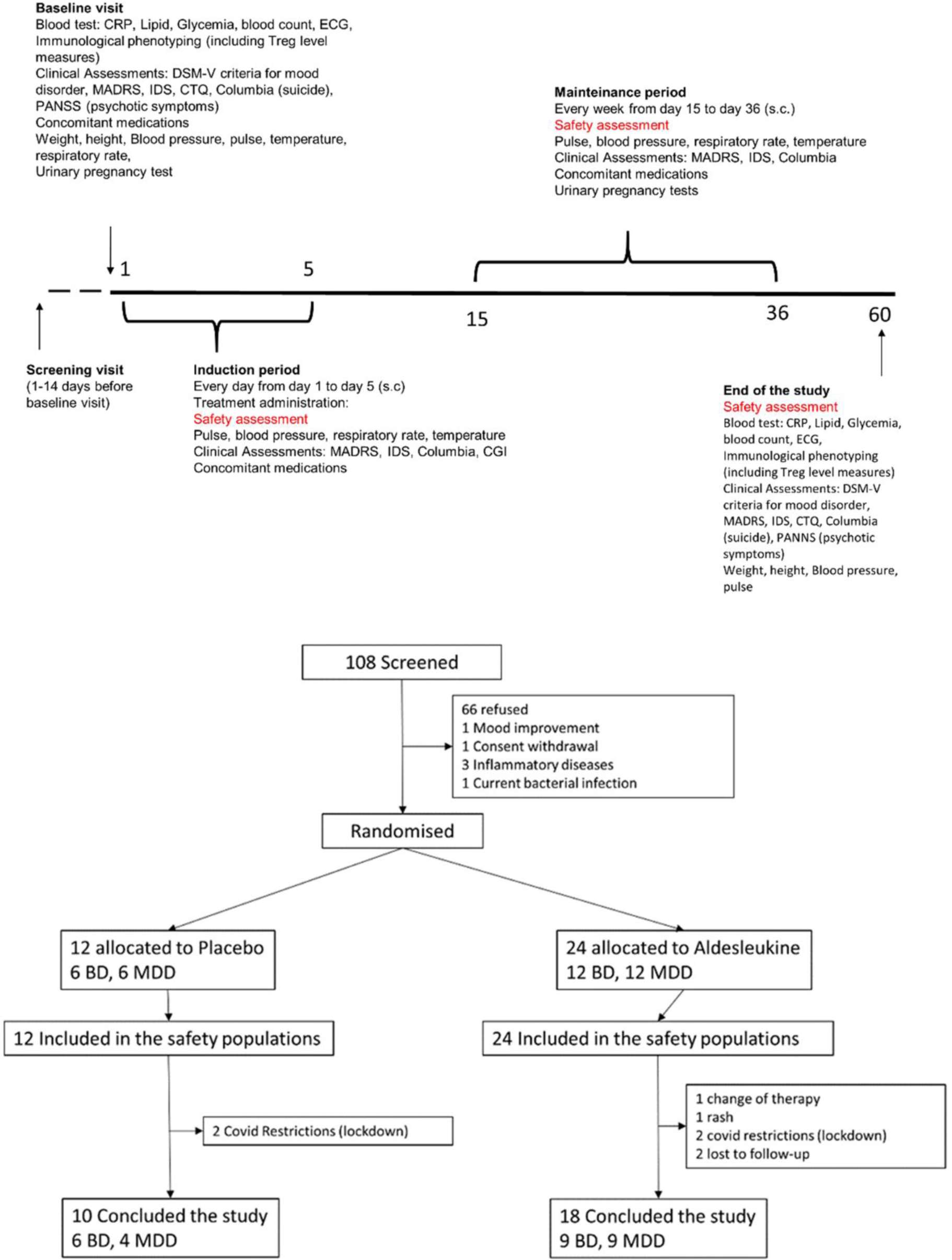
Study design and CONSORT flow diagram.

The study was a single-center, randomised, double-blind, placebo-controlled phase II trial evaluating the safety, clinical efficacy and biological responses of low-dose IL-2 in depressed patients with MDD or BD. IL-2REG was designed and conducted in accordance with the International Conference on Harmonization Good Clinical Practice guidelines and the Declaration of Helsinki, and the study protocol, patient information sheets, and informed consent forms were approved by the Ospedale San Raffaele ethical committee (Prot. N. 77/int/2019) and by the italian regulatory authority (AIFA).

After confirmation of their eligibility at baseline, 36 consecutively recruited inpatients at the Mood Disorder Unit were randomised in a 2:1 ratio to receive either aldesleukin (n=24; 12 MDD and 12 BD) or placebo (n=12; 6 MDD and 6 BD). Treatment randomisation was performed following a randomization list created through the statistical software SPSS. Patients, investigators, nurses, people involved in the evaluation of patients and data managers were kept blind to the treatment allocation for the whole duration of the study and up to the final database lock. The study sponsor remained masked to the individual treatment arm allocation up to the freezing of the database at the end of the study.

Human recombinant IL-2 was provided by the Hospital Pharmacy as Aldesleukin, and administered for 5 weeks as add-on to ongoing antidepressant treatment, with no wash out period. Aldesleukin at a dose of 1 M IU/day or placebo was administered by subcutaneous injection in the morning, every day for consecutive 5 days (induction phase), and then once a week from day 15 to day 36 (maintenance phase). Patients were then followed up until day 60. Dose and scheme of IL-2 administration were in agreement with established protocols for the treatment of inflammatory disorders (Louapre et al., 2023).

All patients were on antidepressant and/or mood stabilizers treatment, and continued it. Treatments had been decided by the psychiatrists in charge, independent from participation to the trial, and kept unchanged. According to our standard treatment protocols, selective serotonin reuptake inhibitors (SSRIs) were preferentially administered; drugs acting on 5-HT and norepinephrine (SNRIs) and tricyclic antidepressants were administered to patients who had not responded to SSRIs in their previous clinical history (https://www.nice.org.uk/guidance/cg90) (Middleton et al., 2005). Patients with BD were also taking lithium.

The first patient was recruited in January 2020, and the last visit was completed in April 2023. Eligible patients were aged 18-65 years, with a diagnosis of MDD or BD (DSM-5 criteria) and an ongoing depressive episode without suicidal ideation. Patients were already on an antidepressant and/or mood stabilizer, and with a Montgomery-Asberg Depression Rating Scale (MADRS) score > 17. Exclusion criteria were: hypersensitivity to active substance or excipient; active infection requiring antibiotics therapy; organ failure (e.g., liver, kidney, lung and heart) or previous history of organ transplantation; immunosuppressive treatment; hepatotoxic, nephrotoxic, myelotoxic or cardiotoxic drugs; any chronic disease; leukocytes<4000/mm^3^, platelets<100 000/mm^3^, hemoglobin<10.0 g/dL, blood red cells <3.5*10^6^/mm^3^; use of anti-inflammatory medication on a regular basis for a chronic inflammatory/autoimmune condition (corticosteroids, NSAID, immunosuppressant IV-Ig); uncontrolled diabetes type I or II; cancer or history of cancer in the last 5 years; existing or planned pregnancy or lactation; for women of child bearing potential, not using a highly effective method of contraception during treatment including oral or injectable contraceptives or intrauterine device or system during participation to the study; immediate risk for suicidal behaviour (3 on HDRS or 5 on MADRS); known HIV infection or clinically manifest AIDS; Parkinson’s or Alzheimer’s disease, or any other serious condition likely to interfere with the conduct of the trial; participation to an interventional study concomitantly or within 30 days prior to this study.

### 2.2 Data collection

The primary endpoint was the change in the relative concentration of peripheral blood Tregs (CD3+CD4+FoxP3+CD127loCD25hi Treg). Secondary endpoints were changes in Th1 and Th2, Naïve T cells, Central memory T cells; changes in a set of peripheral molecules associated with low-grade inflammation in mood disorders; safety and antidepressant efficacy of aldesleukin at day 36 (end of treatment) and at day 60 (end of follow up).

At each visit (day 0-5, 15, 22, 29, 36, 60) severity of depression was rated on the Montgomery-Åsberg Depression Rating Scale (MADRS), Hamilton rating scale for depression (HDRS), Inventory for Depressive Symptomatology Self-Rated (IDS-SR), and safety was assessed based on reported adverse events and changes in concomitant medications. Plasma samples and live peripheral blood mononuclear cells (PBMC) for immunological phenotyping were collected at baseline, day 5, and day 60 (Figure 1).

For immunophenotyping, blood was collected by venipuncture and gradient-purified PBMC were cryopreserved until used. Data were compensated and analysed using FlowJo v.10.8.1 (FlowJo LLC, Ashland, OR). An immune profile was generated on PBMC using a multiparametric flow cytometry. A 28-color flow cytometry panel was used to characterize lymphocyte phenotype and function (panel below). Percentages of viable lymphocytes, T-cells (CD3+) and T cell subpopulation (T-cytotoxic CD8+ and T-helper CD4+) were identified. CD45RA and CCR7 markers were included in the panel to identify naïve and memory T cells. Antibodies that measures T cell function through detection of cytokines (IL-17, IFNγ, TNFα, GM-CSF, IL-2, IL-4) were also included. Percentage of Tregs was assessed (detailed procedure in **Supplementary Methods**).

For measuring peripheral analytes, blood was collected in 6 ml BD Serum tube, increased with silica act clot activator and silicone-coated interior. Patients’ serum was used to quantitative determination respectively of Interleukin 6 (IL-6), Interleukin 7 (IL-7), C-Reactive Protein (CRP), Brain-Derived Neurotrophic Factor (BDNF), Soluble Interleukin 2 Receptor (sIL-2R/sCD25) by means of apDia (Advanced Practical Diagnostics Bv, Turnhout, Belgium) ELISA kits. For each analyte (IL-6, IL-7, CRP, BDNF, sIL-2R/sCD25) an independent ELISA protocol was performed. A standard curve was obtained by plotting the absorbance values versus the corresponding calibrator values. The concentration of the specific analyte was determined by interpolation from the calibration curve (detailed procedure in **Supplementary Methods**).

### 2.3 Statistics

To account for the multiple covarying variables, and considering the *a priori* expected significant interaction with several independent factors (age, sex, diagnosis) and the non-normal distribution of inflammatory biomarkers, we tested the effect of predictors on the outcomes by combinining non-parametric Generalized Linear Model (GLZM) analyses of variances, and linear machine-learning (ML) multiple regression techniques to perform feature reduction and prediction of effects. This robust approach has been shown to succesfully capture the complex relationship between immunological variables and clinical phenotypes and outcomes in the field of mood disorders (Benedetti et al., 2021; Benedetti and Vai, 2023; Mazza et al., 2021; Poletti et al., 2021).

The effect of treatment, sex, and diagnosis was estimated on changes in biomarkers, and on changes in depression severity (delta values baseline-after treatment), by entering independent variables into a GLZM analysis of homogeneity of variances with an identity link function (McCullagh and Nelder, 1989). Parameter estimates were obtained with iterative re-weighted least squares maximum likelihood procedures. The significance of the effects was calculated with the likelihood ratio (LR) statistic, by performing sequential tests for the effects in the model of the factors on the dependent variable, at each step adding an additional effect into the model contributing to incremental χ^2^ statistic, thus providing a test of the increment in the log-likelihood attributable to each current estimated effect; or with the Wald W^2^ test as appropriate (Agresti, 1996; Dobson, 1990).

To assess global response to treatment and perform a feature reduction by selecting the factors of interest in predicting it, we used partial least squares regression (PLS), a ML technique that models the relationships between sets of observed variables with latent variables, to define a linear regression model by projecting the predicted variables and the observable variables to a new space. Changes in rating scales for depression (IDS-SR, MADRS, HDRS) from baseline to day 36 (end of treatment) and day 60 (end of study) were entered as dependent variables, and clinical (treatment, diagnosis, sex) and biological (delta cell counts and levels of analytes) variables were entered in the model as predictors. Accuracy and significance of the predictive value of the model was assessed by using the Nonlinear Iterative Partial Least Squares (NIPALS) algorithm (Wold, 1966) and optimizing by cross-validation the number of PLS components to extract (A), then calculating R^2^X (a A-dimensional vector, to record the explained variance of the data matrix of predictors by each PLS component), R^2^Y (a A-dimensional vector, to record the explained variance of response variables by each PLS component), Q^2^ (predicted variation, to measure R^2^Y applied to a test set with cross-validation procedure, in order to assess the predictive relevance of the endogenous constructs); and for each variable predictive weights (w), and the variable importance in projection (VIP) values, to estimate the contribute of each variable to the explanation of Y variance and the direction of effect. K-fold cross-validation was performed by randomly splitting the data into k folds of roughly equal size, in order to estimate the error rate of the predictive algorithm by resampling the analysis data on k-1 folds to then assess the performance on the final fold. Significance of the model and of variable contributions were defined by Q^2^>0 and VIP>1 (Akarachantachote et al., 2014; Chong and Jun, 2005; Hill and Lewicki, 2006; Palermo et al., 2009). Given the preliminary nature of the study, a cutoff value at VIP>0.8 for mining variables potentially contributing to prediction was however considered (SAS Institute, 2017).

All the statistical analyses were performed with a commercially available software package (StatSoft Statistica 12, Tulsa, OK, USA) and following standard computational procedures (Dobson, 1990; Hill and Lewicki, 2006).

## 3. Results

Clinical and demographic characteristics of the participants are resumed in **Table 1**. Between January 2020 and April 2023, 108 patients were screened for eligibility, 36 were enrolled, and 28 patients concluded the study, thus allowing to assess the primary endpoint (**Figure 1**). All patients met criteria for TRD (Sforzini et al., 2022) except 3 patients with BD, 2 treated with placebo and 1 with aldesleukin.

**Table 1.** Clinical and demographic characteristics of the patients, and levels of significance of the differences based on treatment options. Values are mean±SD.

| All enrolled patients |  |  |  |  |  |  |  |  |  |  |  |  |
| --- | --- | --- | --- | --- | --- | --- | --- | --- | --- | --- | --- | --- |
|  | Whole sample n=36 |  |  |  | MDD n=18 |  |  |  | BD n=18 |  |  |  |
| | Placebo n=12 | IL-2 n=24 | t/$\chi^2$ | p | Placebo n=6 | IL-2 n=12 | t/$\chi^2$ | p | Placebo n=6 | IL-2 n=12 | t/$\chi^2$ | p |
| Age | 48.23±15.76 | 52.12±10.46 | 0.901 | 0.373 | 46.17±18.82 | 50.53±13.78 | 0.573 | 0.573 | 50.00±13.91 | 54.00±4.12 | 0.906 | 0.378 |
| Sex | 9F 4M | 14F 10M | 0.425 | 0.514 | 5F 1M | 7F 5M | 0.902 | 0.342 | 3F 3M | 6F 6M | 0.011 | 0.913 |
| Onset | 24.69±8.55 | 33.92±10.52 | 2.707 | 0.010 | 25.16±12.28 | 35.92±12.12 | 1.790 | 0.573 | 24.28±4.49 | 31.54±8.16 | 2.139 | 0.048 |
| Duration of illness | 23.53±13.12 | 17.70±9.84 | 1.528 | 0.135 | 21.00±12.96 | 14.61±10.29 | 1.160 | 0.261 | 25.71±13.87 | 21.36±8.27 | 0.839 | 0.413 |
| N of depressive episodes | 5.75±13.12 | 7.34±7.66 | 0.670 | 0.507 | 4.83±2.85 | 8.38±9.79 | 0.859 | 0.402 | 6.67±5.24 | 6.00±3.46 | 0.308 | 0.762 |
| Duration of episode | 20.46±20.95 | 13.82±16.71 | 1.043 | 0.303 | 22.00±17.62 | 15.25±12.41 | 0.947 | 0.357 | 19.14±24.78 | 12.27±20.97 | 0.632 | 0.536 |
| Body Mass Index | 26.28±5.07 | 24.48±3.77 | 1.224 | 0.228 | 25.74±7.24 | 24.03±4.61 | 0.629 | 0.537 | 26.75±2.70 | 25.03±2.56 | 1.360 | 0.192 |
| MADRS | 31.61±7.47 | 32.87±7.52 | 0.487 | 0.629 | 31.33±5.92 | 33.69±9.04 | 0.579 | 0.570 | 31.85±9.08 | 31.90±5.46 | 0.015 | 0.988 |
| HDRS | 22.76±4.16 | 24.50±5.21 | 1.029 | 0.310 | 22.50±1.37 | 25.00±5.55 | 1.072 | 0.298 | 23.00±5.74 | 23.90±4.98 | 0.355 | 0.726 |
| IDS-SR | 36.56±12.44 | 38.54±12.51 | 0.465 | 0.644 | 41.00±7.77 | 37.00±10.59 | 0.823 | 0.421 | 32.71±14.91 | 40.36±14.80 | 1.065 | 0.302 |
| SSRIs | 6 (50%) | 14 (61%) | 0.503 | 0.477 | 3 (50%) | 6 (50%) | 0.024 | 0.875 | 3 (50%) | 8 (%) | 1.606 | 0.205 |
| SNRI | 3 (50%) | 7 (28%) | 0.158 | 0.690 | 2 (50%) | 6 (50%) | 0.276 | 0.598 | 1 (%) | 1 (%) | 0.116 | 0.732 |
| Antipsychotics | 6 (50%) | 11 (50%) | 0.216 | 0.641 | 2 (50%) | 6 (50%) | 0.276 | 0.598 | 5 (%) | 5 (%) | 1.168 | 0.279 |
| Mood stabilizer | 10 (80%) | 15 (72) | 0.800 | 0.370 | 3 (50%) | 4 (44%) | 0.652 | 0.419 | 6 (100%) | 12 (100%) | - | - |
| Patients who completed the treatment |  |  |  |  |  |  |  |  |  |  |  |  |
|  | Whole sample n=28 |  |  |  | MDD n=13 |  |  |  | BD n=15 |  |  |  |
| | Placebo n=10 | IL-2 n=18 | t/$\chi^2$ | p | Placebo n=4 | IL-2 n=9 | t/$\chi^2$ | p | Placebo n=6 | IL-2 n=9 | t/$\chi^2$ | p |
| Age | 44.9±16.64 | 52.5±11.19 | 1.445 | 0.160 | 40.00±20.91 | 51.89±15.81 | 1.140 | 0.278 | 48.17±14.29 | 53.11±3.91 | 1.000 | 0.335 |
| Sex | 6F 4M | 10F 8M | 0.051 | 0.819 | 3F 1M | 6F 3M | 0.090 | 0.763 | 3F 3M | 4F 5M | 0.044 | 0.832 |
| Onset | 25.1±9.70 | 32.94±10.09 | 1.997 | 0.056 | 25.25±15.77 | 34.89±11.51 | 1.251 | 0.237 | 25.00±4.47 | 31.00±8.67 | 1.549 | 0.145 |
| Duration of illness | 19.8±12.58 | 18.89±9.99 | 0.210 | 0.834 | 14.75±11.11 | 17.00±11.07 | 0.337 | 0.741 | 23.17±13.28 | 20.78±9.01 | 0.417 | 0.683 |
| N of depressive episodes | 5±4.24 | 8.76±8.47 | 1.244 | 0.225 | 3.75±1.70 | 10.89±10.95 | 1.265 | 0.231 | 6.00±5.57 | 6.37±3.81 | 0.145 | 0.887 |
| Duration of episode | 24.4±22.58 | 15.47±18.79 | 1.106 | 0.279 | 29.00±17.93 | 16.87±14.21 | 1.283 | 0.228 | 21.33±26.40 | 14.22±22.93 | 0.554 | 0.588 |
| BMI | 27.27±5.43 | 25.07±3.87 | 1.250 | 0.222 | 27.13±8.89 | 24.99±4.88 | 0.572 | 0.578 | 27.37±2.36 | 25.15±2.83 | 1.581 | 0.137 |
| MADRS | 30.80±7.84 | 31.44±6.91 | 0.225 | 0.823 | 31.25±6.70 | 30.44±8.07 | 0.173 | 0.865 | 30.50±9.13 | 32.44±5.83 | 0.506 | 0.620 |
| HAMD | 22.40±4.57 | 23.17±4.75 | 0.414 | 0.682 | 22.50±1.73 | 22.89±4.51 | 0.484 | 0.673 | 22.33±5.99 | 23.44±5.24 | 0.158 | 0.709 |
| SSRIs | 5 (50%) | 11 (61%) | 0.324 | 0.569 | 2 (50%) | 3 (33%) | 0.325 | 0.568 | 3 (50%) | 8 (89%) | 2.784 | 0.095 |
| SNRI | 3 (30%) | 5 (28%) | 0.015 | 0.900 | 2 (50%) | 5 (56%) | 0.034 | 0.853 | 1 (16%) | 0 | 1.607 | 0.204 |
| Antipsychotics | 6 (60%) | 9 (50%) | 0.258 | 0.611 | 2 (50%) | 4 (44%) | 0.034 | 0.852 | 4 (67%) | 5 (56%) | 0.185 | 0.667 |
| Mood stabilizer | 8 (80%) | 13 (72) | 0.207 | 0.648 | 2 (50%) | 4 (44%) | 0.034 | 0.852 | 6 (100%) | 9 (100%) | - | - |

All patients completed the induction phase. Treatment was generally well tolerated and no serious adverse reactions (SARs) not serious adverse events (SAEs) were observed. Only transient and mild events were observed, the most frequent being injection site reactions, with two patients showing a mild allergic reaction with a rapid resolution without treatment, and 1 patient with a mild allergic reaction who resolved with an antihistaminic treatment (thus excluded from the study). Drop-outs occurred because of direct effect of COVID restrictions (lockdown) in 4 patients, with 2 additional patients lost for not willing to come to the hospital during the pandemic. One patients necessitated a change in antidepressant drug treatments. All drop-outs except one occurred before the end of the treatment phase. Patients who completed the trial did not significantly differ in administered combined antidepressant drugs.

### 3.1 Biological outcomes

Treatment influenced immune cell counts (**Table 2**). Patterns of change of Treg and CD4+ and CD8+ Naïve cell counts did not follow parallel slopes of time course in the two treatment groups (**Figure 2**).

**Figure 2.**
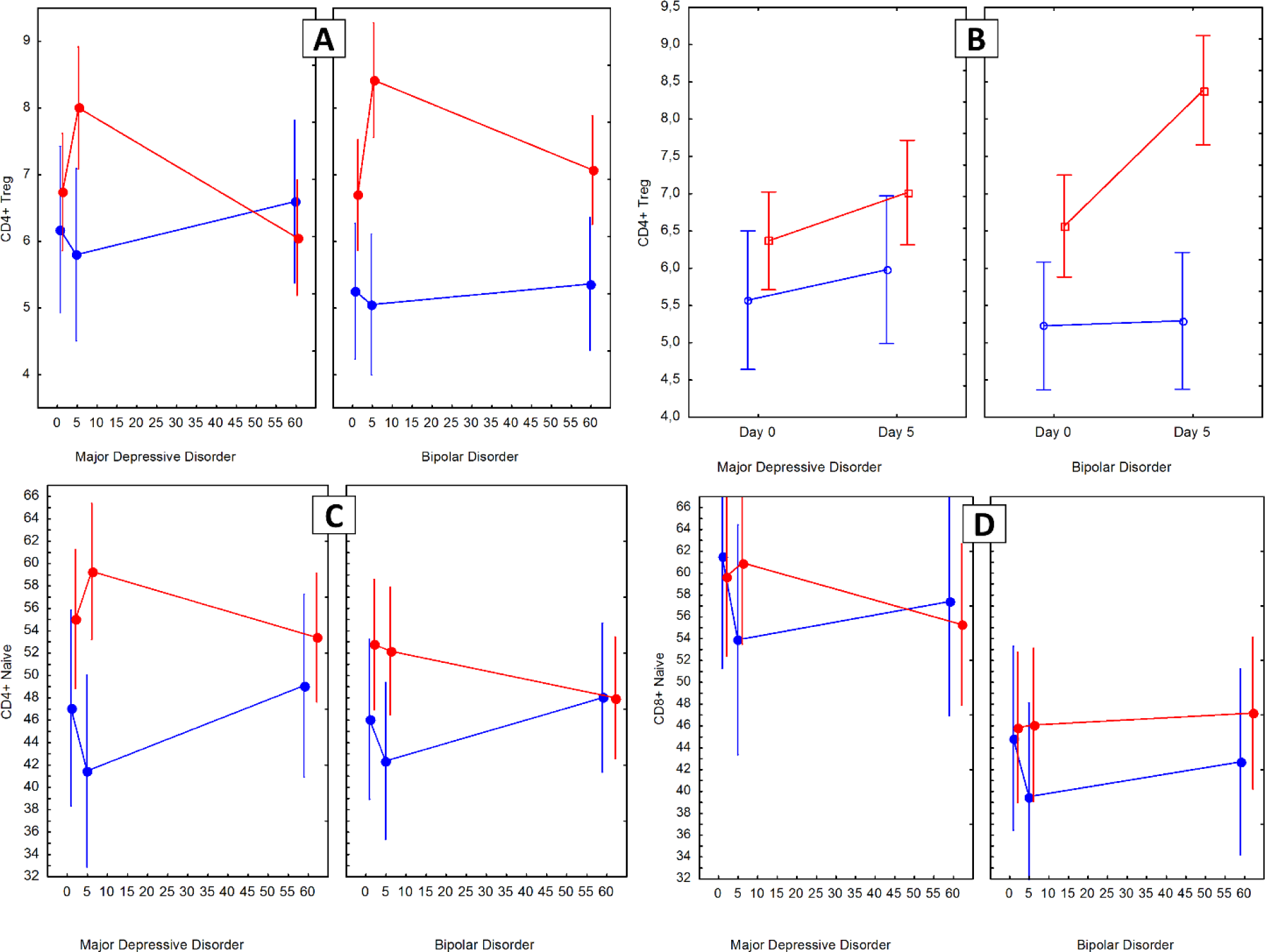
Changes in CD4+ Treg cell counts in participants who completed the trial (A, n=28) or who completed the induction phase (B, n=36). Changes in CD4+ Naive (C), CD8+ Naive (D) cell counts during treatment (frequencies of CD4+ and CD8+). Red=Aldesleukin; Blue=Placebo. Points are means, whiskers are SEM.

**Table 2.**
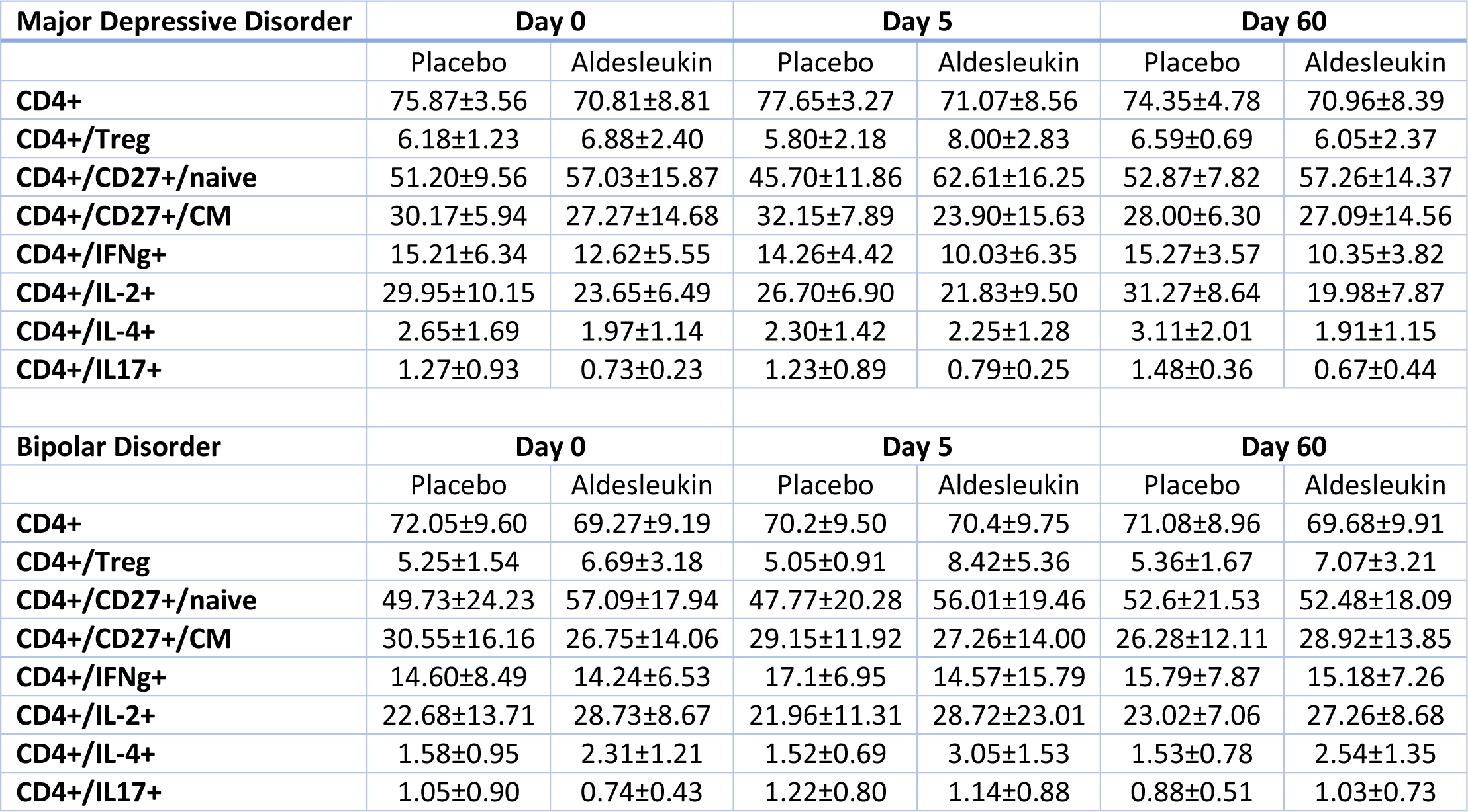
Changes of cell counts (frequency of CD4+) during treatment.

| Major Depressive Disorder | Day 0 |  | Day 5 |  | Day 60 |  |
| --- | --- | --- | --- | --- | --- | --- |
|  | Placebo | Aldesleukin | Placebo | Aldesleukin | Placebo | Aldesleukin |
| CD4+ | 75.87±3.56 | 70.81±8.81 | 77.65±3.27 | 71.07±8.56 | 74.35±4.78 | 70.96±8.39 |
| CD4+/Treg | 6.18±1.23 | 6.88±2.40 | 5.80±2.18 | 8.00±2.83 | 6.59±0.69 | 6.05±2.37 |
| CD4+/CD27+/naive | 51.20±9.56 | 57.03±15.87 | 45.70±11.86 | 62.61±16.25 | 52.87±7.82 | 57.26±14.37 |
| CD4+/CD27+/CM | 30.17±5.94 | 27.27±14.68 | 32.15±7.89 | 23.90±15.63 | 28.00±6.30 | 27.09±14.56 |
| CD4+/IFNg+ | 15.21±6.34 | 12.62±5.55 | 14.26±4.42 | 10.03±6.35 | 15.27±3.57 | 10.35±3.82 |
| CD4+/IL-2+ | 29.95±10.15 | 23.65±6.49 | 26.70±6.90 | 21.83±9.50 | 31.27±8.64 | 19.98±7.87 |
| CD4+/IL-4+ | 2.65±1.69 | 1.97±1.14 | 2.30±1.42 | 2.25±1.28 | 3.11±2.01 | 1.91±1.15 |
| CD4+/IL17+ | 1.27±0.93 | 0.73±0.23 | 1.23±0.89 | 0.79±0.25 | 1.48±0.36 | 0.67±0.44 |
| Bipolar Disorder | Day 0 |  | Day 5 |  | Day 60 |  |
|  | Placebo | Aldesleukin | Placebo | Aldesleukin | Placebo | Aldesleukin |
| CD4+ | 72.05±9.60 | 69.27±9.19 | 70.2±9.50 | 70.4±9.75 | 71.08±8.96 | 69.68±9.91 |
| CD4+/Treg | 5.25±1.54 | 6.69±3.18 | 5.05±0.91 | 8.42±5.36 | 5.36±1.67 | 7.07±3.21 |
| CD4+/CD27+/naive | 49.73±24.23 | 57.09±17.94 | 47.77±20.28 | 56.01±19.46 | 52.6±21.53 | 52.48±18.09 |
| CD4+/CD27+/CM | 30.55±16.16 | 26.75±14.06 | 29.15±11.92 | 27.26±14.00 | 26.28±12.11 | 28.92±13.85 |
| CD4+/IFNg+ | 14.60±8.49 | 14.24±6.53 | 17.1±6.95 | 14.57±15.79 | 15.79±7.87 | 15.18±7.26 |
| CD4+/IL-2+ | 22.68±13.71 | 28.73±8.67 | 21.96±11.31 | 28.72±23.01 | 23.02±7.06 | 27.26±8.68 |
| CD4+/IL-4+ | 1.58±0.95 | 2.31±1.21 | 1.52±0.69 | 3.05±1.53 | 1.53±0.78 | 2.54±1.35 |
| CD4+/IL17+ | 1.05±0.90 | 0.74±0.43 | 1.22±0.80 | 1.14±0.88 | 0.88±0.51 | 1.03±0.73 |

Treatment with aldesleukin, but not with placebo, caused a significant expansion of Treg cells at the end of the induction phase (day 5), as confirmed by a GLZM homogeneity of slopes analysis showing a significant effect of treatment on Delta day0-day5 Treg cell counts (LR χ^2^=4.603, p=0.0320) with no main effects nor interactions with age, sex, and diagnosis in the 28 participants who completed the trial. The effect was not anymore apparent at the end of the follow-up (day 60), when cell counts did not anymore significantly differ from baseline levels. Considering all patients who had completed the induction phase (n=36), significant interactions were detected: for the effect of treatment with diagnosis (higher effect of aldesleukin in patients with BD, W^2^=4.127, p=0.0422); for the effect of age, which was positively associated with an increase in Treg cell counts (W^2^=6.841, p=0.0090) after placebo, but not after aldesleukin, which caused a higher increase independent of age (Treatment x Group x Age interaction: W^2^=4.865, p=0.0274) (**Supplementary** Figure 1).

CD4+ Naïve cells increased with aldesleukin in MDD patients but not in BD patients, while they decreased in placebo treated patients, yielding a significant effect of treatment in the whole group (LR χ^2^=6.241, p=0.0125) on day0-day5 cell counts, which was not mantained over time. Similar effects were observed for CD8+ Naïve cells, which were higher in MDD at baseline, decreased in placebo-treated patients, but remained sustantially stable in aldesleukin treated patients, again showing a significant treatment effect on day0-day5 changes (LR χ^2^=11.115, p=0.0009).

With an opposite trend (**Supplementary Figure 2**), central memory CD4+ CD27+ CM cell counts showed a decrease with aldesleukin and an increase with placebo after the induction phase in MDD, but not in BD patients, who showed an opposite trend persisting at Day 60. This yielded a significant Treatment x Diagnosis interaction (LR χ^2^=5.273, p=0.0217), with no effect of treatment alone in the whole sample. Similar effects were observed for CD8+ CD27+ CM cell counts (treatment effect in the whole group: W^2^=4.100, p=0.0430). These effects led to an increase in the CD4+ Naïve/CM ratio (W^2^=4.208, p=0.0402), and to a nominal, but non-significant increase in CD8+ Naïve/CM ratio.

Changes of Th1, Th2, and Th17 cell counts are shown in **Figure 3**. Th17 cell counts showed not significant changes in the studied groups, with a trend to increse in all groups except MDD patients treated with placebo. CD4+ IFNγ+ counts (Th1) decreased in MDD, and increased or remained stable in BD during the induction phase, with a significant effect of diagnosis on delta day0-day5 (LR χ^2^=5.277, p=0.0216) and no effect of treatment. CD4+ IL-4+ cell counts (Th2) increased during the induction phase in patients treated with aldesleukin, but not with placebo, with a significant effect of treatment (LR χ^2^=7.963, p=0.0048) and not of diagnosis.

**Figure 3.**
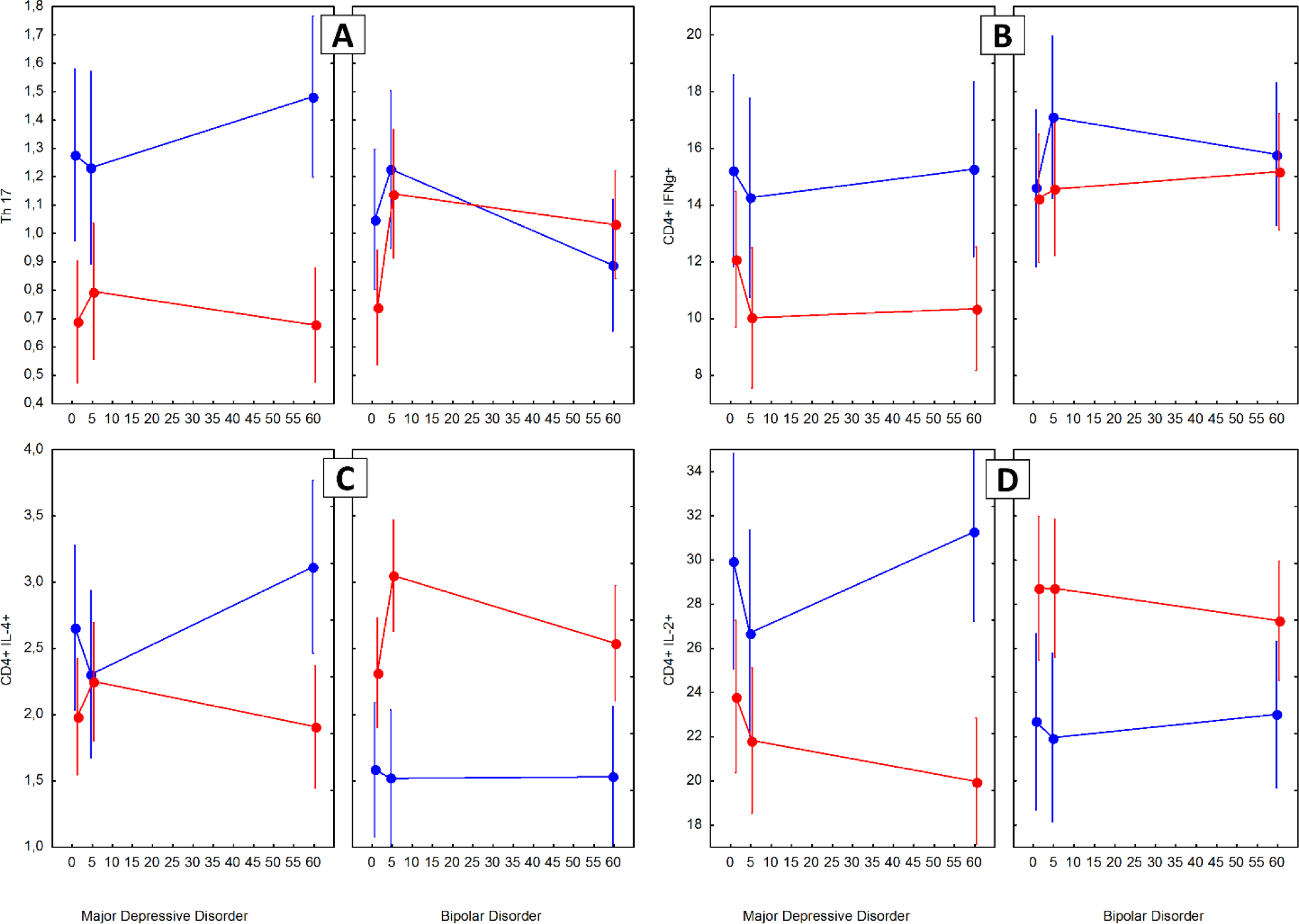
Changes in Th17 (A), CD4+ IFNγ+ (B), CD4+ IL-4+ (C), and CD4+ IL-2+ (D) cell counts during treatment (frequencies of CD4+). Red=Aldesleukin; Blue=Placebo. Points are means, whiskers are SEM.

Finally, levels of sCD25 (sIL-2Rα) significantly increased during the induction phase in MDD patients treated with aldesleukin, and in BD patients irrespective of treatment options, yielding a significant effect of treatment in the whole sample (LR χ^2^=11.115, p=0.0009). CRP showed a significantly higher increase after aldesleukin during the induction phase (LR χ^2^=9.749, p=0.0018). Aldesleukin did not significantly affect levels of BDNF and IL-7 (**Table 3, Supplementary Figure 3**). Baseline levels of IL-6 were below the sensitivity limits in 17/28 participants, and in 4/28 at the end of the induction phase, with a significant increase (Wilcoxon Z=2.915, p=0.0036) independent of treatment.

**Table 3.**
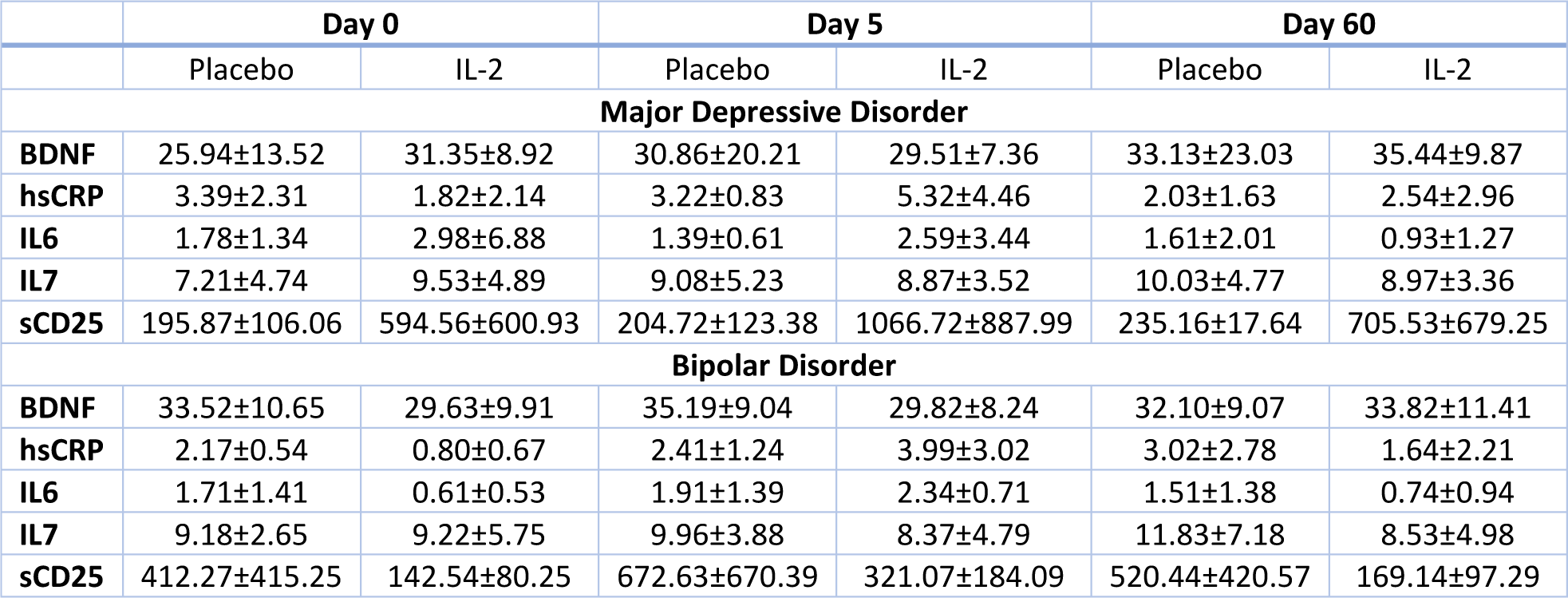
Changes of inflammatory biomarkers during treatment (pg/ml).

### 3.2 Antidepressant efficacy of low dose IL-2 treatment

Aldesleukin add-on treatment was followed by a significantly better amelioration of depression severity than placebo. Inspection of patterns of change in severity of depression (**Figure 4**), and of improvement (delta scores) from baseline to the end of treatment (Day 36) and to the end of follow-up (Day 60) (**Figure 5**) show erratic changes during the first weeks, and then an advantage for low-dose IL-2 over placebo.

**Figure 4.**
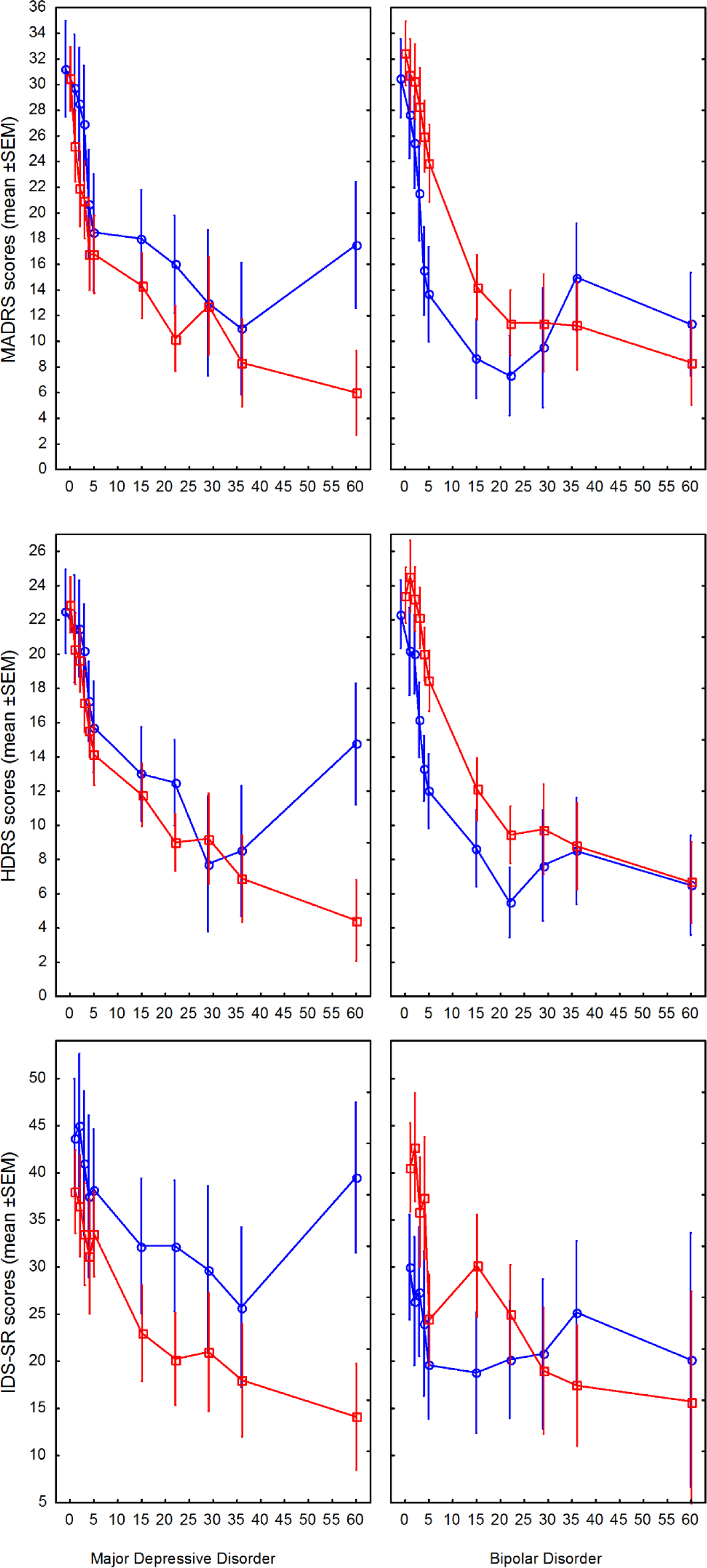
Pattern of change of severity of depression as rated at MADRS (top), HDRS (middle), and IDS-SR (bottom) during treatment and follow-up. Red=Aldesleukin; Blue=Placebo. Points are means, whiskers are SEM.

**Figure 5.**
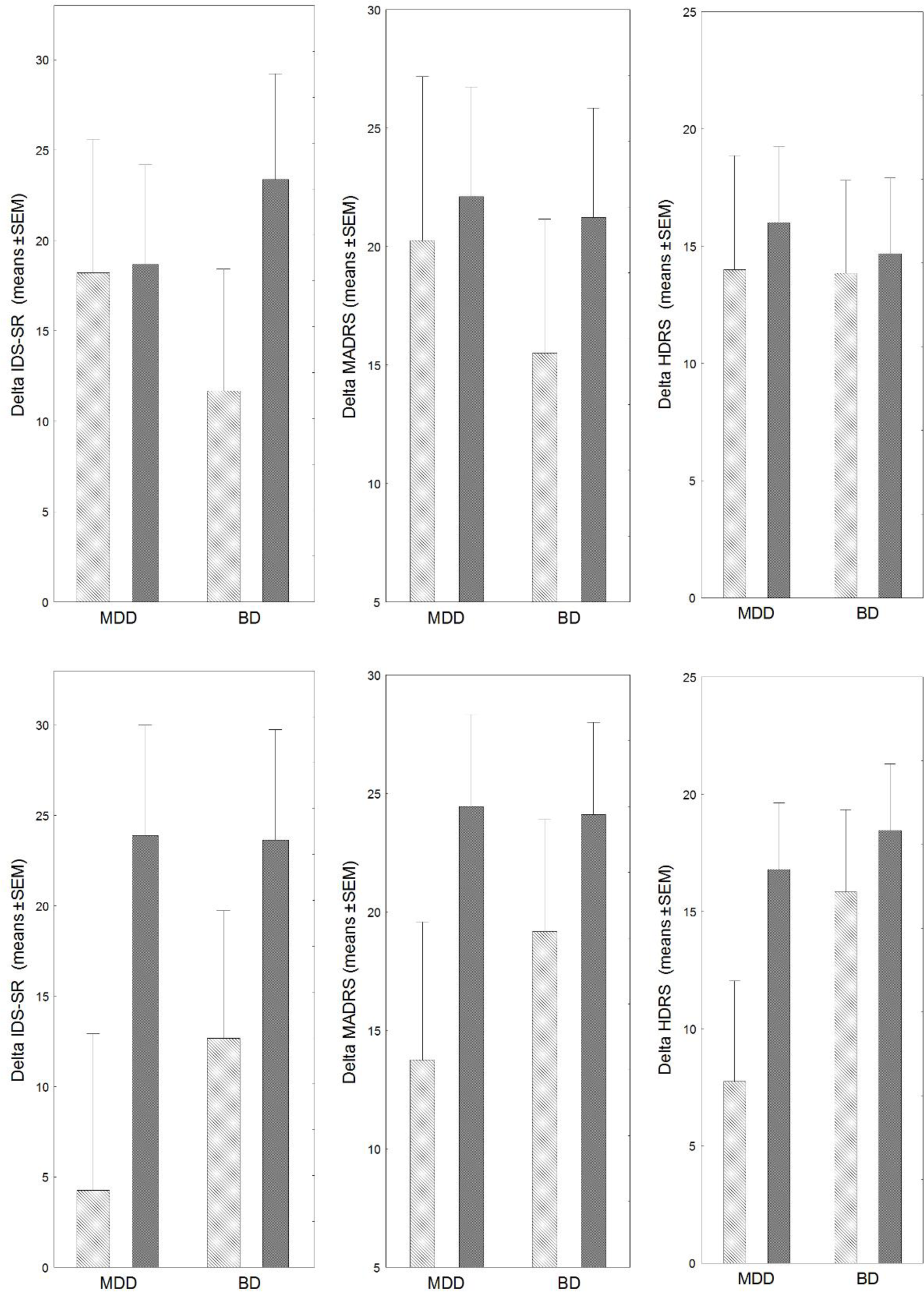
Improvement in severity of depression as rated at MADRS (top), HDRS (middle), and IDS-SR (bottom) during treatment (Top, from baseline to Day 36) and follow-up (Bottom, from baseline to Day 60). Pale grey= Placebo; Dark grey= Aldesleukin. Bars are means, whiskers are SEM.

A PLS linear regression with treatment, diagnosis, sex, and age as predictors of the global improvement at Day 60 defined a significant model predicting the efficacy of treatment when considering all the rated dimensions of depression (IDS-SR, MADRS, HDRS), with one significant component (coefficient=0.562) explaining 19% of variance in improvement: R²X=0.300; R²Y=0.185; Q²=0.031. The variable selected as relevant to predict outcomes were treatment (aldeslukin vs placebo) (VIP=1.37) and age (VIP=1.61), while sex (VIP=0.47) and diagnosis (VIP=0.29) were dismissed as not relevant.

Using an explanatory approach on single dimensions of improvement, separated GLZM homogeneity of slopes ANOVAs showed a significant effect of treatment both at day 36 and at day 60, and on all rated dimensions of depression severity. At day 36, treatment significantly affected changes in MADRS (W^2^=4.723, p=0.0298), HDRS (W^2^=8.365, p=0.0038), and IDS-SR (W^2^=6.367, p=0.0116) scores, with a significant interaction with Diagnosis at HDRS (better effect in MDD, W^2^=7.002, p=0.0081). At day 60, treatment again significantly affected changes in MADRS (W^2^=6.571, p=0.0104), HDRS (W^2^=9.359, p=0.0022), and IDS-SR (W^2^=8.094, p=0.0044) scores, with a significant effect of diagnosis (lower scores in MDD) at MADRS (W^2^=5.016, p=0.0251) and HDRS (W^2^=7.699, p=0.0055), but no significant Treatment x Diagnosis interaction, meaning (in agreement with PLS results) that the benefit from aldeslukin versus placebo was independent from diagnosis.

Age significantly correlated with self-rated improvement at IDS-SR at day 36 (r=0.84, p=0.002) and at day 60 (r=0.85, p=0.002) and with improvement at HDRS at day 36 (r=0.75, p=0.013) in the placebo treated group, but not in the aldesleukin treated group. With multivariate GLZM modelling, significant Treatment x Age interactions were observed at all rating scales both, at day 36 (MADRS: W^2^=5.691, p=0.0017; HDRS: W^2^=10.839, p=0.0010; IDS-SR: W^2^=7.182, p=0.0074), and at day 60 (MADRS: W^2^=5.945, p=0.0148; HDRS: W^2^=8.504, p=0.0035; IDS-SR: W^2^=6.887, p=0.0087).

### 3.3 Biological predictors of antidepressant efficacy

Effects of treatment on immune cell counts during the induction phase predicted the subsequent improvement in severity of depression.

With a ML predictive approach, the previously described linear PLS regression model was enriched with the changes induced by treatment before/after the induction phase (Day 0-5) in the *a priori* defined biomarkers (primary endpoint of the study) as predictors: changes in immune cell composition and activation status (CD4+ TRegs, CD4+ Naïve, CD8+ Naïve, CD4+ IL-17+, CD4+ IFN+, CD4+ IL-2+, CD4+ IL-4+, CD4+ CM, CD8+ CM cell counts), and circulating levels of hsCRP, sCD25, IL-6, IL-7, and BDNF.

Global improvement at day 60, considering all the rated dimensions of depression (IDS-SR, MADRS, HDRS), was significantly predicted by a model with one significant component explaining 33.5% of variance in depression improvement (coefficient=0.497, R²X=0.213; R²Y=0.335; Q²=0.094) (**Figure 6**). The variables significantly contributing to predict outcomes were Treatment (w=0.298, VIP=1.312), Age (w=0.329, VIP=1.326), and delta changes of CD4+ Naïve (w=0.409, VIP=1.655), CD4+ IL-4+ (w=-0.380, VIP=1.577), CD4+ Tregs (w=0.096, VIP=1.386), CD8+ Naïve (w=0.294, VIP=1.251), CD4+ IL-17+ (w=0.036, VIP=1.194), and hsCRP (w=345, VIP=1.458). Changes in CM cell counts were marginally relevant (CD4+: w=0.231, VIP=0.926; CD8+: w=0.202, VIP=0.861). Direction of the effect for delta changes was positive (i.e., an increase of cell counts associated with better improvement) for all variables except for CD4+ IL-17+, CD4+ IFN+, and levels of IL-6 and IL-7, for which a decrease associated with better improvement.

**Figure 6.**
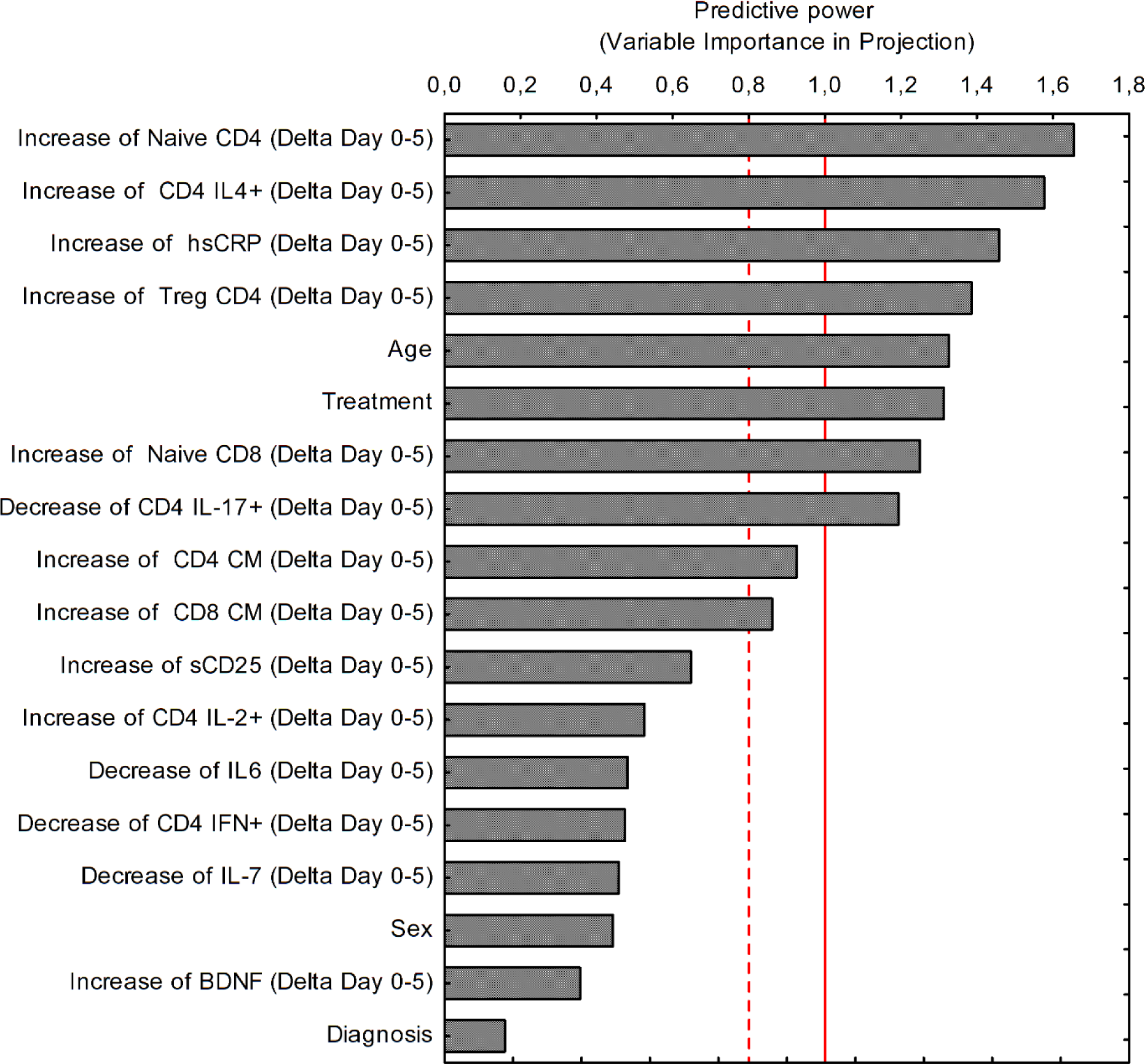
Variables predicting the global improvement in depression severity at study end (Day 60), as rated at MADRS, HDRS, and IDS-SR during treatment, listed according to their relative predictive power at linear regression; and direction of the effect contributing to improvement (increase vs decrease).

The same ML analysis performed on depression improvement at day 36 (end of IL-2 treatment) selected 5 biomarkers whose changes significantly contributed to predict the improvement: delta changes of CD4+ Naïve (w=0.431, VIP=1.973), CD4+/IL-4+ (w=0.337, VIP=1.545), CD8+ CM (w=0.422, VIP=1.936), CD4+ CM (w=0.346, VIP=1.586), with a marginal contribution of CD4+ IL-17+ (w=0.203, VIP=0.931). A model considering only these biomarkers showed a significant effect, explaining 31.3% of variance in outcomes at day 36 (coefficient=0.624, R²X=0.440; R²Y=0.313; Q²=0.158).

Post-hoc, separated GLZM homogeneity of slopes ANOVAs performed on single outcomes (delta improvement in severity at rating scales) at the two times, using the features selected as relevant by the linear PLS regression (delta changes in biomarkers during the induction phase), confirmed the statistical significance of delta CD4+ Tregs and delta CD4+ Naïve cell counts in predicting outcomes, yielding significant differential effects of the other biomarkers at different times and outcomes (**Table 4**).

**Table 4.** Levels of significance of the effects of biological predictors of improvement (delta changes during the induction phase, Day0-Day5) on the improvement in severity of depression at Day 36 and at Day 60, as measured on MADRS, HDRS, and IDS-SR.

|  | CD4+ Naïve | CD4+ IL-4+ | CD4+ TRegs | CD8+ Naïve | CD4+ IL-17+ | hsCRP | CD4+ CM | CD8+ CM |
| --- | --- | --- | --- | --- | --- | --- | --- | --- |
| **Day 36** |  |  |  |  |  |  |  |  |
| **MADRS** | $W^2=4.466$<br>$p=0.0346$ | $W^2=5.940$<br>$p=0.0148$ | NS | NS | $W^2=5.367$<br>$p=0.0205$ | NS | $W^2=4.260$<br>$p=0.0390$ | $W^2=5.320$<br>$p=0.0211$ |
| **HDRS** | $W^2=6.084$<br>$p=0.0136$ | NS | $W^2=4.047$<br>$p=0.0442$ | $W^2=3.875$<br>$p=0.0490$ | NS | $W^2=4.702$<br>$p=0.0301$ | NS | NS |
| **IDS-SR** | NS | $W^2=11.701$<br>$p=0.0006$ | NS | NS | NS | NS | NS | NS |
| **Day 60** |  |  |  |  |  |  |  |  |
| **MADRS** | $W^2=5.141$<br>$p=0.0234$ | $W^2=20.401$<br>$p<0.0001$ | $W^2=12.394$<br>$p=0.0004$ | NS | NS | $W^2=6.848$<br>$p=0.0089$ | NS | NS |
| **HDRS** | $W^2=18.082$<br>$p<0.0001$ | NS | $W^2=19.428$<br>$p<0.0001$ | NS | NS | NS | NS | $W^2=5.741$<br>$p=0.0166$ |
| **IDS-SR** | $W^2=39.598$<br>$p<0.0001$ | NS | $W^2=9.515$<br>$p=0.0020$ | $W^2=10.954$<br>$p=0.0009$ | NS | $W^2=7.126$<br>$p=0.0076$ | NS | NS |

## 4. Discussion

We observed significant effects of treatment with low dose IL-2 vs placebo in expanding the population of Treg, Th2, and Naive CD4+/CD8+ immune cell counts, and in potentiating treatment efficacy when added-on to ongoing antidepressant drugs. Changes in cell counts were rapidly induced in the first five days of treatment, and predicted the later improvement of depression severity.

Associating immune changes with the clinical depression improvement in the weeks following the induction phase, and persisting in the month after the end of treatment, speaks of complex relationships between the immune system and the clinical phenotype of mood disorders. The pattern of variation of self- and observer rating scales for depression showed a consistent trend toward amelioration in the first month of treatment, followed by a trend to relapse in the second month, consistent with current consensus about clinical outcomes of TRD, where a very high relapse rate is expected while continuing treatment even if it was apparently effective at the beginning (Sforzini et al., 2022). Counteracting this trend, well evident in placebo-treated patients, the main effect of IL-2 was to enhance response at 5 weeks of treatment and prevent the subsequent relapse, thus promoting and consolidating improvement over time.

Several non-alternative biological mechanisms could contribute to the observed antidepressant effect of IL-2.

IL-2 is a T cell growth factor, which in low dose can particularly increase the cell counts of Tregs, an effect repeatedly reported in human trials in immune diseases and associated with clinical benefits (see Introduction). Here this effect predicted the antidepressant potentiation. Moreover, IL-2 increased counts of Th2 cells, which support the intrinsic anti-inflammatory properties of the brain (Gimsa et al., 2001), and again this effect predicted antidepressant improvement. In a previous report we found Treg frequencies inversely associated with the pro-inflammatory state of monocytes (Grosse et al., 2016), and it is generally assumed that high Treg cell frequencies correlate with reduced inflammation. However, the possible anti-inflammatory correlate of this effect was not revealed by CRP and IL-6, which have been considered promising baseline markers of inflammation associated with major depression (Arteaga-Henriquez et al., 2019; Pitharouli et al., 2021). Counterintuitively, these markers of low grade inflammation increased with low dose IL-2 treatment and also predicted its efficacy. Interestingly, such an effect has been earlier observed after SSRIs/SNRIs antidepressant drugs, proportional to reduction in depression severity (Carboni et al., 2019; Hannestad et al., 2011).

Due to its effect on Treg cells, low dose IL-2 treatment is also expected to decrease the Th17/Treg ratio, which is higher in depression, and has been proposed as a hallmark of severity and suicidality in MDD, and as a target for antidepressant treatment (Cui et al., 2021; Schiweck et al., 2022). In our study Th17 cells tended to an average increase with low dose IL-2 treatment, but within this average pattern of change, it was a relative negative trend (to a decrease or a lower increase) which predicted better final antidepressant improvement. This observation suggests that Th17 cells might have complex interactions with the depressed brain. This is also illustrated by the reports that Th17 cells are essential for hippocampal neuroplasticity (Niebling et al., 2014), and that high levels of Th17 cells correlated significantly to better integrity brain white matter (WM) in the of BD patients and healthy controls (Poletti et al., 2017). Further research is needed to clarify their role in mood disorders.

IL-2 is also known to increase thymic production of naïve CD4+ T cells (Carcelain et al., 2003), and it indeed increased CD4+ and CD8+ Naïve cells in this trial, both effects predicting antidepressant improvement. A reduction of naïve T cells and an expansion of memory and senescent T cells is a core characteristics of the immunological imbalance associated with mood disorders, associating with abnormally activated monocytic/macrophagic innate immunity setpoints (Simon et al., 2021b). The imbalance between lower naïve, and higher memory T cells was reported to be proportional to severity of MDD, also characterizing suicide risk (Schiweck et al., 2020). Changes of all these cell populations counts are then surmised to promote a general rebalancing of the innate/adaptive immunity imbalance in mood disorders.

Interestingly, IL-2 may also act as a trophic factor on both neurons and oligodendrocytes (de Araujo et al., 2009). Studies on neurons and astrocytes consistently showed that IL-2 promoted survival and neurite extension of cultured cortical, hippocampal, septal, striatal, and cerebellar neurons (Hanisch and Quirion, 1995). *In vivo* animal models of neuroinflammatory-neurodegenerative disorders showed induction of astrocytic activation and improved synaptic plasticity and spine density in the hippocampus (Alves et al., 2016), also promoting hippocampal neurogenesis (Liu et al., 2014). Reduced hippocampal volumes are one of the most robust findings in brain imaging studies of depressed patients (Schmaal et al., 2016), and also among the few consistent brain structural predictors of poor treatment response (Enneking et al., 2020). We recently showed that lower hippocampal volume predicted worse response to SSRIs and SNRIs administered upon clinical need in a hospital setting (Paolini et al., 2023a), and that it partially mediated the detrimental effect of peripheral low-grade inflammation on antidepressant response (Paolini et al., 2023b). It can then be surmised that the neurotrophic effect of IL-2 could contribute to correct these effects.

Mood disorders associate with signs of disrupted brain WM integrity, which are influenced by inflammatory markers, and correlate with severity and outcome of the disease (Benedetti et al., 2016; Benedetti et al., 2011; Favre et al., 2019; Van Velzen et al., 2020). Human oligodendrocytes express the IL-2 receptor, and while *in vitro* at higher concentration a combined IL-1/IL-2 administration hampered oligodendrocyte progenitor cell proliferation (Saneto et al., 1986), at low concentration IL-2 alone markedly stimulated the proliferation of normal human oligodendrocytes (Otero and Merrill, 1997). IL-2 could also promote myelin regeneration by stimulating Treg cells, which promoted oligodendrocyte progenitor cell differentiation and myelination in vitro (Dombrowski et al., 2017). We also showed that *in vivo* the Th17 and Treg relative frequencies play key roles in myelin maintenance and regeneration, with higher Th17 associated with higher fractional anisotropy in the core WM skeleton of the brain (Poletti et al., 2017). WM integrity predicts antidepressant response both in MDD and in BD (Bollettini et al., 2015; Gerlach et al., 2022). Rapid antidepressant response associates with a rapid improvement in WM integrity (Melloni et al., 2020). Low-dose IL-2 expanded both, Treg and Th17 in the present trial: it can be surmised that the combined direct and indirect effects of IL-2 on WM integrity could contribute to correct the abnormalities hampering antidepressant response.

The trophic effects of IL-2 on brain cells are likely to be mediated by the IL-2 activation of the PI3 kinase (PI3K) activity, an upstream regulator of glycogen synthase kinase 3-β (GSK-3β), acting via Akt to phopsphorylate ser9 and leading to inactivation of GSK-3β (Braunstein et al., 2008). GSK-3β is involved in the control of gene expression, cell behavior, cell adhesion, neuronal polarity, and in regulation of neurodevelopment, neuronal plasticity and cell survival (Grimes and Jope, 2001). Increased GSK-3β activity was reported in *post mortem* brain tissue of depressed suicides, and GSK-3β inhibition is a unique common feature of mood stabilizers lithium and valproate, serotonergic antidepressants, and some antipsychotics of proven efficacy in BD (Beaulieu et al., 2009; Li and Jope, 2010). In turn, in animal models the GSK-3β inhibitor lithium prolonged T cell proliferation and increased IL-2 production (Ohteki et al., 2000), and the serotonergic antidepressant fluoxetine increased IL-2 (Fazzino et al., 2009) also restoring T cell proliferation and IL-2 levels after chronic stress-driven immune system depression (Frick et al., 2009). It can then be hypothesized that IL-2, also promoted by lithium and antidepressant treatment, could be synergistic with ongoing treatment in rescuing the impaired neuroplasticity associated with mood disorders (Machado-Vieira et al., 2013) by promoting cortical and hippocampal neuroplasticity as influenced by GSK-3β (Manji et al., 2003), thus paving the way to antidepressant response.

Strengths of the present study include a focused research question and state-of-the-art methods, but our results must be viewed in light of some limitations. The COVID pandemic occurred during the study, limiting access to the hospital infrastructures and directly increasing attrition. No patient was drug-naive, and the drug treatments administered during the course of the illness and of the current episode could have influenced biological outcomes. Recruitment was in a single center and in a single ethnic group, thus raising the possibility of population stratifications.

These limitations, however, do not bias the main finding of an effect of low-dose IL-2 in modulating the immune system and in promoting antidepressant response in patients with MDD and BD, thus providing the first RCT evidence supporting the hypothesis that treatment to strengthen the T cell system could be a successful way to correct the immuno-inflammatory abnormalities associated with mood disorders, and potentiate antidepressant response. Interest is warranted for combining multimodal approaches and techniques from computational, molecular and biological psychiatry to deepen the understanding of the immune-brain interaction, disentangle the intertwined signaling pathways which trigger and shape the phenotype of the disorder and its outcome, and define predictors and correlates of response to cluster patients for a precision, personalized treatment approach.

## Data Availability

All data produced in the present study are available upon reasonable request to the authors

## Acknowledgments

The authors thanks Cristian Beccaria and Matteo Iannacone (Division of Immunology, Transplantation and Infectious Diseases, IRCCS San Raffaele Scientific Institute, Milan, Italy), and Luca Battistini (Neuroimmunology Unit, IRCCS Santa Lucia Foundation, Rome, Italy) for the precious help in immunophenotyping.

The design of the IL2REG and DEPIL-2 trials stemmed from a joint effort by FB, SP, RF in Milano; and Marion Leboyer (Université Paris Est Créteil, INSERM U955, IMRB, Laboratoire Neuro-Psychiatrie translationnelle, Créteil), David Klatzmann (Sorbonne Université-INSERM UMRS959, Immunology-Immunopathology-Immunotherapy laboratory, Paris), and their collaborators; prompted and coordinated by HAD (https://moodstratification.eu/).

## Declarations of interest: **none**

