## Supplementary material for "Low-dose interleukin 2 antidepressant potentiation in unipolar and bipolar depression: Safety, efficacy, and immunological biomarkers"

#### Supplementary materials

### 1. Supplementary methods

#### 1.1 FACS immunophenotyping gating strategy

For immunophenotyping, blood was collected by venipuncture and gradient-purified PBMC were cryopreserved until used. Peripheral blood mononuclear cells (PBMCs) were rapidly thawed and cells were rested in 10% FBS-RPMI at 37°C in a humidified 5% CO2 incubator for 2 h before stimulation. Viability was assessed by 0.4% trypan blue dye exclusion. $1x10^6$ cells were seeded in a U-bottom 96-well plate and stimulated with PMA (30ng/ml) and Ionomycin (1000 ng/ml) for 4h at 37°C. Brefeldin A was added (1mg/ml) to the culture for the last 2 hours of stimulation. Post-culture cells were pelleted in a V-bottom 96-well plate and resuspended in 50 µL of surface monoclonal antibodies at pre-optimized concentrations and diluted in Brilliant Stain Buffer (BD Biosciences), then incubated for 20' at RT in the dark. For intracellular staining, cells were fixed and permeabillized for 30' at 4°C using Foxp3 / Transcription Factor Staining Buffer Set (eBioscience™). Cells were then incubated for 30' in 50 µL 1X Permeabilization Buffer with the recommended amount of directly conjugated antibody for detection of intracellular antigens. Samples were acquired on a fully equipped Cytek Aurora within 24 hours from staining and fixation. Data were compensated and analysed using FlowJo v.10.8.1 (FlowJo LLC, Ashland, OR). An immune profile was generated on PBMC using a multiparametric flow cytometry. A 28-color flow cytometry panel was used to characterize lymphocyte phenotype and function (panel below). Percentages of viable lymphocytes, T-cells (CD3+) and T cell subpopulation (T-cytotoxic CD8+ and T-helper CD4+) were identified. CD45RA and CCR7 markers were included in the panel to identify naïve and memory T cells. Antibodies that measures T cell function through detection of cytokines (IL-17, IFNγ, TNFα, GM-CSF, IL-2, IL-4) were also included. Additionally, percentage of Tregs was assessed.

| Fluorochrome | Antigen | Company |
|---|---|---|
| FITC | Granzyme B | BioLegend |
| BB630 | CD57 | BD Horizon |
| PercPCy5.5 | CD69 | BD Pharminger |
| BB700 | CD28 | BD Optibuild |
| BB790-P | TNFa | BD Horizon |
| PE | IL-4 | BD Pharminger |
| PECF594 | GM-CSF | BD Horizon |
| PE-Cy5 | CD56 | Immunotech |
| PE-Cy5.5 | FoxP3 | Invitrogen |
| PeCy7 | CCR7 | BioLegend |
| APC | IFNg | BD Pharminger |
| cFluor R720 | Perforin | Cytek |
| APC-Cy7 | CD45RA | BioLegend |
| BV421 | IL17 | BioLegend |
| BV510 | Live Dead<br>CD19<br>CD14 | Thermo Fisher<br>Biolegend<br>Biolegend |
| BV570 | TCRVα7.2 | BD Horizon |
| BV605 | CD161 | BioLegend |
| BV650 | CD49d | BD Optibuild |
| BV711 | KLRG1 | BioLegend |
| BV750 | TCR γδ2 | BD Optibuild |
| BV786 | CD27 | BD Horizon |
| BUV395 | CD3 | BD Horizon |
| BUV496 | CD4 | BD Horizon |
| BUV563 | CD25 | BD Horizon |
| BUV615 | CD39 | BD Optibuild |
| BUV661 | PD-1 | Bd Optibuild |
| BUV737 | IL-2 | BD Horizon |
| BUV805 | CD8 | BD Horizon |

| Emission range (nm) | UltraViolet 355 | | | Violet 405 | | | Blue 488 | | | Yellow Green 561 | | | Red 640 | | |
|---|---|---|---|---|---|---|---|---|---|---|---|---|---|---|---|
| | Det | Fluorochrome | Target | Det | Fluorochrome | Target | Det | Fluorochrome | Target | Det | Fluorochrome | Target | Det | Fluorochrome | Target |
| 365-380 | UV1 | | | | | | | | | | | | | | |
| 380-395 | UV2 | BUV395 | CD3 | | | | | | | | | | | | |
| 420-435 | UV3 | | | V1 | BV421 | IL-17 | | | | | | | | | |
| 435-450 | UV4 | | | V2 | | | | | | | | | | | |
| 450-465 | UV5 | | | V3 | | | | | | | | | | | |
| 465-480 | UV6 | | | V4 | | | | | | | | | | | |
| 498-518 | UV7 | BUV496 | CD4 | V5 | | | B1 | | | | | | | | |
| 516-533 | | | | V6 | | | B2 | FITC | GrzB | | | | | | |
| 528-556 | UV8 | | | V7 | BV510 | LD/CD19/CD14 | B3 | | | | | | | | |
| 571-590 | UV9 | BUV563 | CD25 | V8 | BV570 | TCRa7.2 | B4 | | | YG1 | PE | IL-4 | | | |
| 588-608 | | | | V9 | | | B5 | | | YG2 | | | | | |
| 605-628 | UV10 | BUV615 | CD39 | V10 | BV605 | CD161 | B6 | BB630 | CD57 | YG3 | PE-CF594 | GM-CSF | | | |
| 650-669 | UV11 | BUV661 | PD-1 | V11 | BV650 | CD49d | B7 | | | YG4 | | | R1 | APC | IFNg |
| 669-687 | | | | | | | B8 | PerCP.Cy5.5 | CD69 | YG5 | PE-Cy5 | CD56 | R2 | | |
| 677-705 | UV12 | | | V12 | | | B9 | BB700 | CD28 | YG6 | | | R3 | | |
| 705-727 | UV13 | | | V13 | BV711 | KLRG1 | B10 | | | YG7 | PE-Cy5.5 | Foxp3 | R4 | cFluor R720 | Perforin |
| 728-749 | | | | | | | B11 | | | | | | R5 | | |
| 735-765 | UV14 | BUV737 | IL-2 | V14 | BV750 | TCR γδ2 | B12 | | | YG8 | | | R6 | | |
| 765-795 | UV15 | | | V15 | BV786 | CD27 | B13 | | | YG9 | Pe-Cy7 | CCR7 | R7 | APC.Cy7 | CD45RA |
| 795-829 | UV16 | BUV805 | CD8 | V16 | | | B14 | BB790 | TNF | YG10 | | | R8 | | |

| | |
|---|---|
| (blue) | Viability |
| (green) | Surface |
| (orange) | Intra |

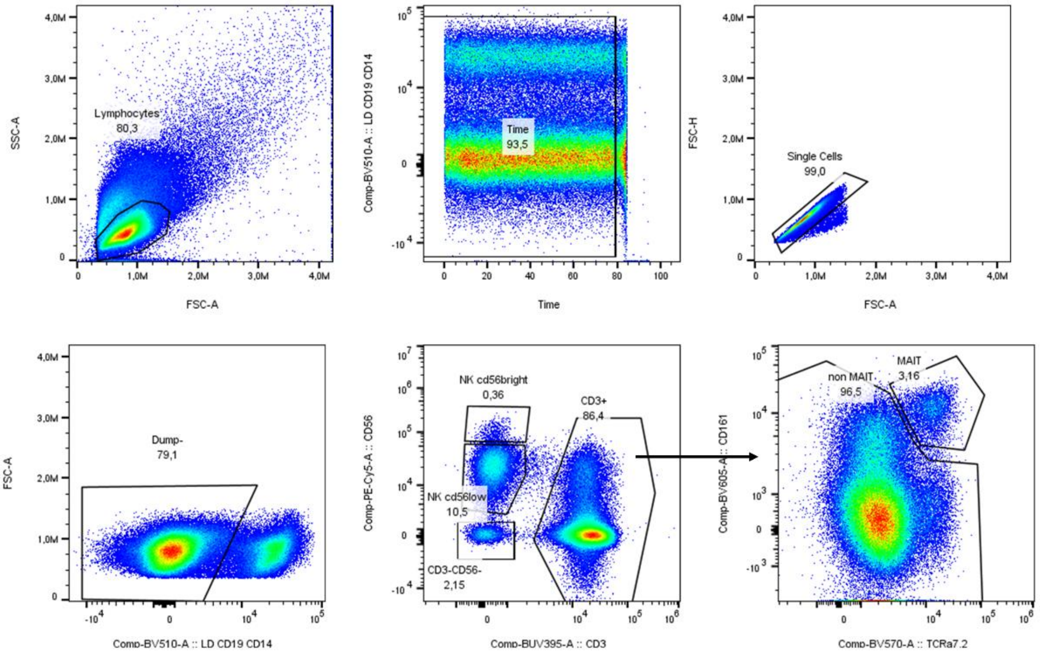


1

Gated on MAIT

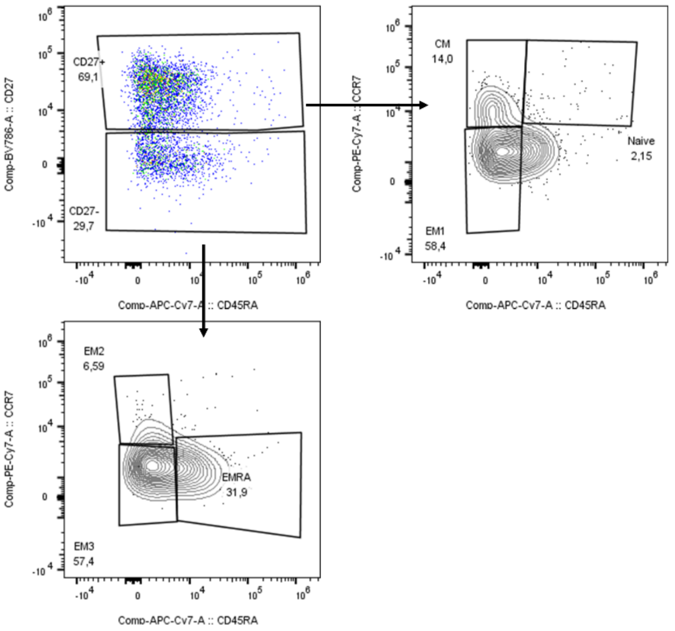


2

Gated on non MAIT

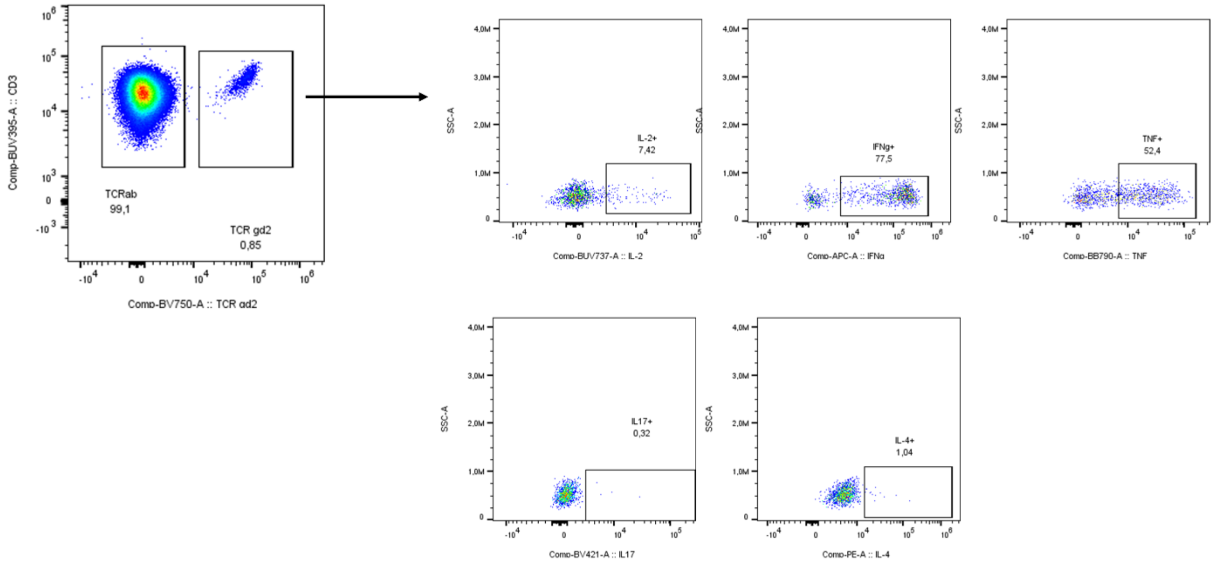


3

Gated on TCRab

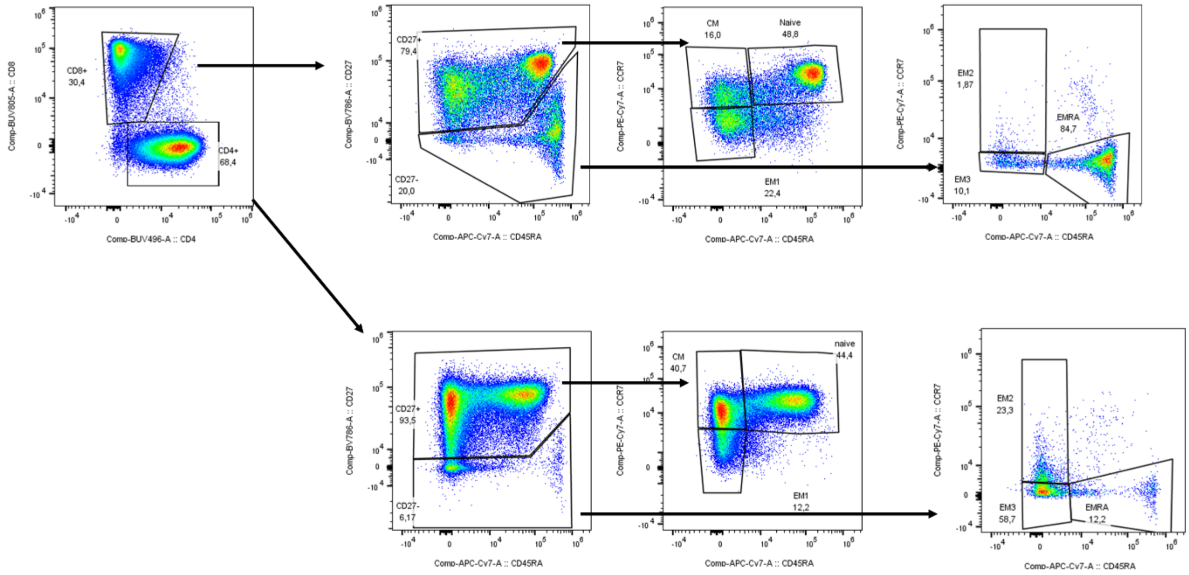


4

Gated on CD4

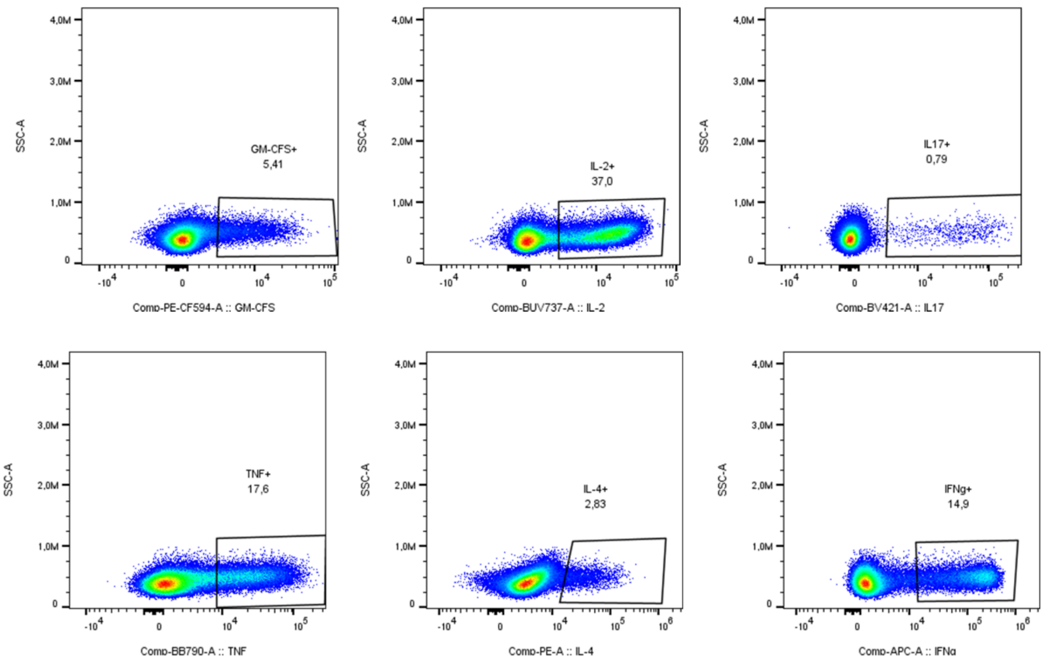


5

Gated on CD4

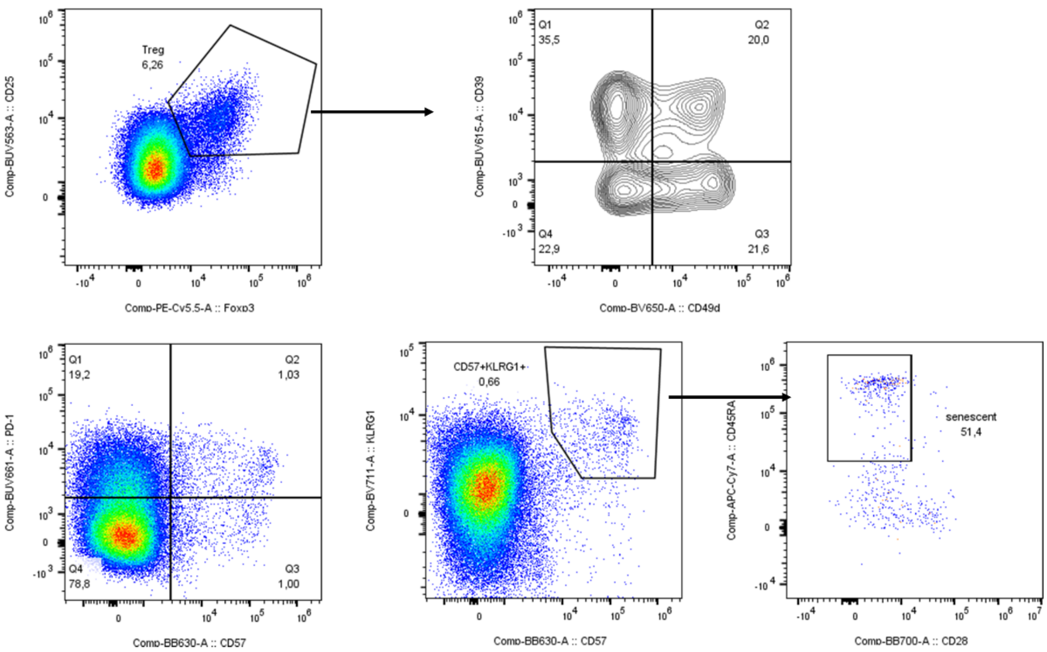


6

Gated on CD8

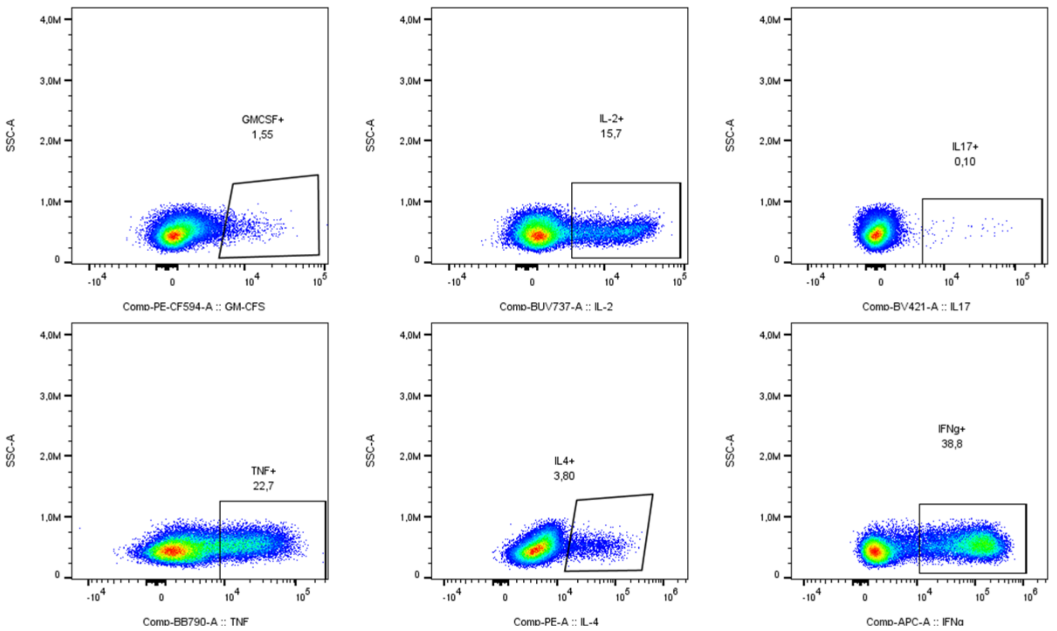


7

Gated on CD8

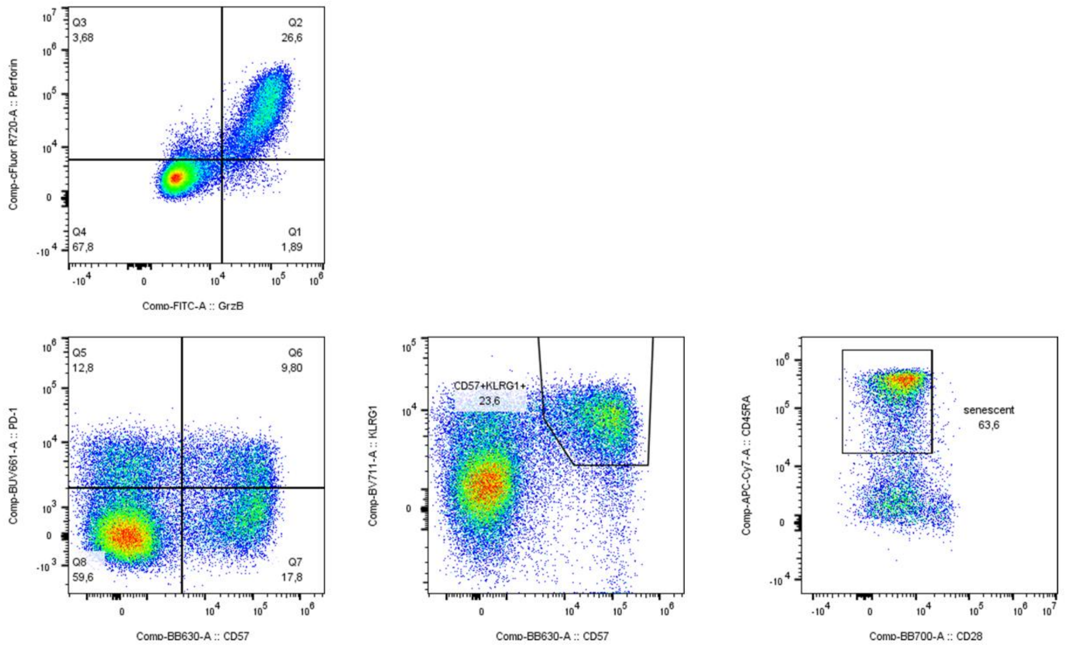


8

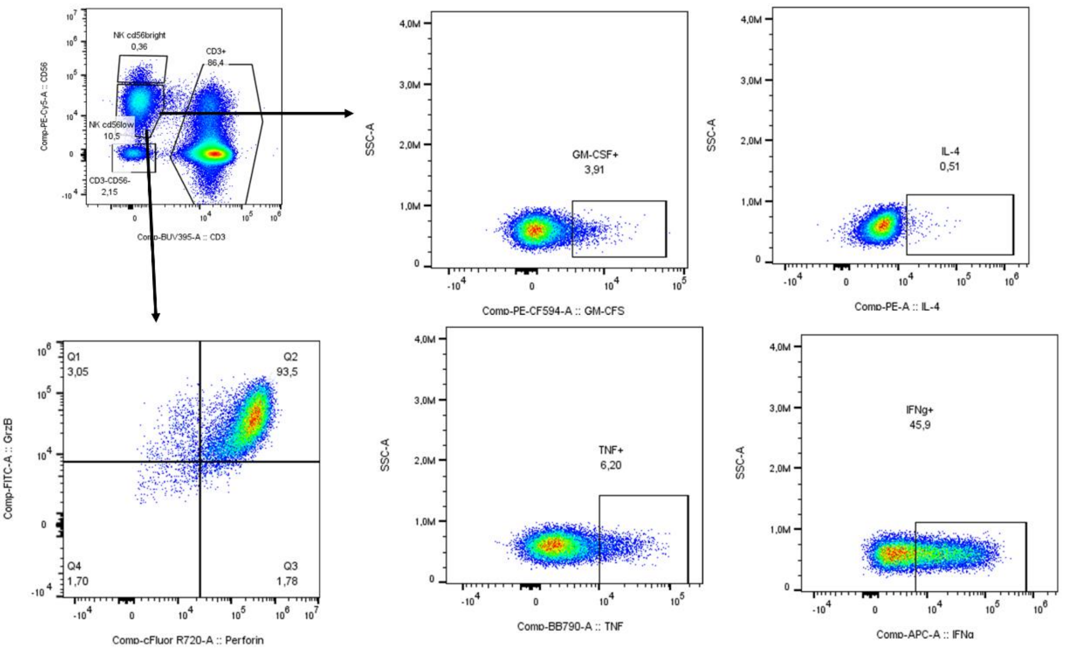
NK cd56bright
0,36
CD3+
86,4
NK cd56low
10,5
CD3-CD56-
2,15
Comp-PE-Cy5-A :: CD56
Comp-BUV395-A :: CD3
GM-CSF+
3,91
Comp-PE-CF594-A :: GM-CFS
SSC-A
IL-4
0,51
Comp-PE-A :: IL-4
Q1
3,05
Q2
93,5
Q4
1,70
Q3
1,78
Comp-FITC-A :: GrzB
Comp-cFluor R720-A :: Perforin
TNF+
6,20
Comp-BB790-A :: TNF
IFNg+
45,9
Comp-APC-A :: IFNg

9

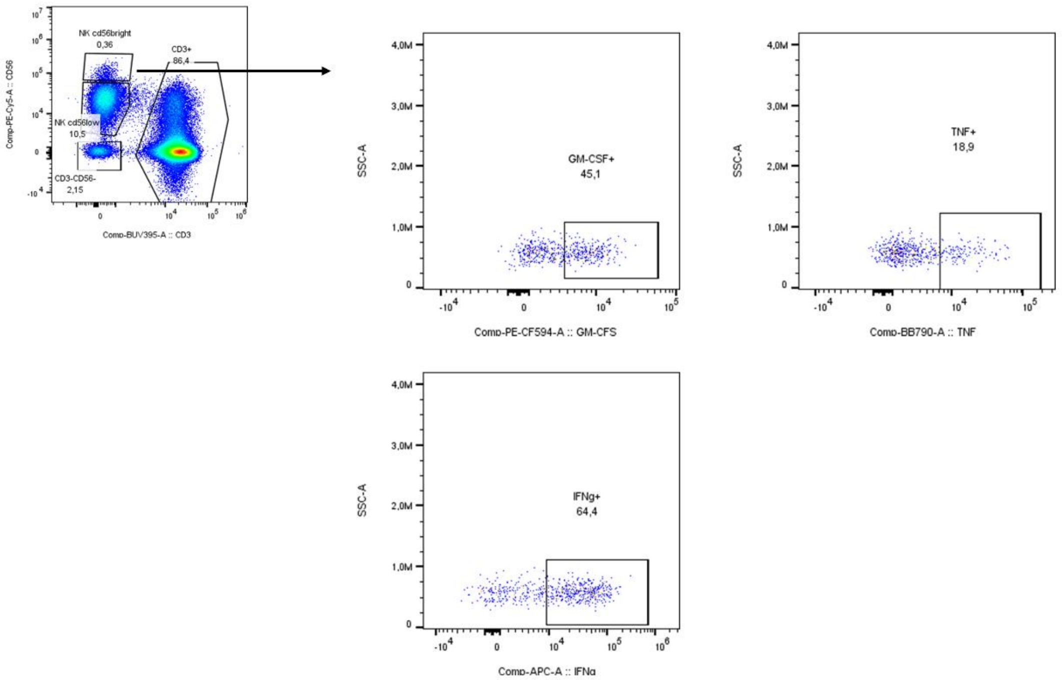
NK cd56bright
0,36
CD3+
86,4
NK cd56low
10,5
CD3-CD56-
2,15
Comp-PE-Cy5-A :: CD56
Comp-BUV395-A :: CD3
GM-CSF+
45,1
SSC-A
Comp-PE-CF594-A :: GM-CFS
TNF+
18,9
Comp-BB790-A :: TNF
IFNg+
64,4
Comp-APC-A :: IFNg

10

#### 2. ELISA assessment of peripheral analytes

For measuring peripheral analytes, blood was collected in 6 ml BD Serum tube, increased with silica act clot activator and silicone-coated interior (BD, Ref 367815). The tube was left for, at least, 30 minutes at room temperature to allow it to clot. The clot was removed by centrifuging at 2500 x g for 10 minutes in a swing out rotor. The supernatant (about 3 mL) was subdivided into 0.2 mL aliquots (Tubes, 0.5 mL, Thermofisher, Ref AB0350) and store at -60°C. Patients' serum was used to quantitative determination respectively of Interleukin 6 (IL-6), Interleukin 7 (IL-7), C-Reactive Protein (CRP), Brain-Derived Neurotrophic Factor (BDNF), Soluble Interleukin 2 Receptor (sIL-2R/sCD25) by means of apDia (Advanced Practical Diagnostics Bv, Turnhout, Belgium) ELISA kits. For each analyte (IL-6, IL-7, CRP, BDNF, sIL-2R/sCD25) an independent ELISA protocol was performed as follow. Microtiter strips, coated with specific monoclonal anti-human antibodies, were incubated with calibrators, a positive control and diluted samples. During this incubation step, specific human antigen binds specifically to the antibodies on the solid phase. After removal of the unbound serum proteins by a washing procedure, the strips were incubated with specific biotin conjugated polyclonal antibodies, binding directly to the antigen-antibody complex. After removal of the unbound biotin conjugate, the strips were incubated with peroxidase-conjugated streptavidin. After removal of the unbound peroxidase conjugate, the strips were incubated with chromogenic solution containing tetramethylbenzidin and hydrogen peroxide: a blue colour develops in proportion to the amount of immunocomplex bound to the wells of the strips. The enzymatic reaction was stopped by the addition of 0.5 M sulfuric acid (H2SO4) and the absorbance values at 450 nm were determined. A standard curve was obtained by plotting the absorbance values versus the corresponding calibrator values. The concentration of the specific analyte was determined by interpolation from the calibration curve.

#### 2. Supplementary results

**Supplementary Figure 1**. Age was positively associated with an increase in Treg CD4+ cell counts ($W^2$=6.841, p=0.0090), but with an effect driven by the strong relationship observed in patients treated with placebo (older age, higher increase in cell counts), but not in patients treated with aldesleukin, who showed a much higher increase, independent of age (Treatment x Group x Age interaction: $W^2$=4.865, p=0.0274).

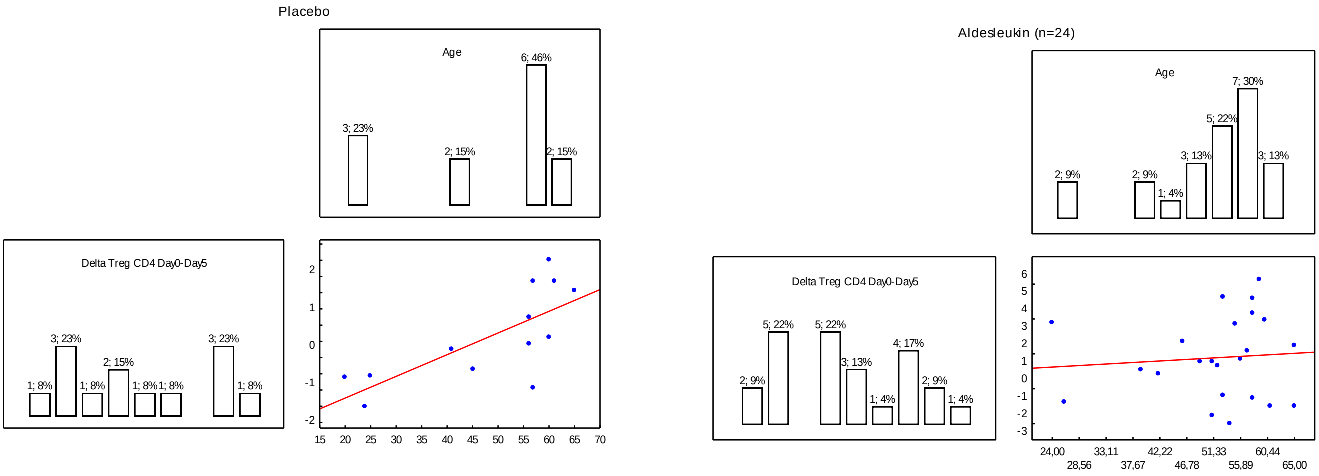

**Supplementary Figure 2**.

Changes in CD4+ CD27+ CM (A), and CD8+ CD27+ CM (B) cell counts during treatment (frequencies of CD4+ and CD8+), and in the CD4+ Naïve/CM and CD8+ Naïve/CM ratio (C and D). Red=Aldesleukin; Blue=Placebo. Points are means, whiskers are SEM.

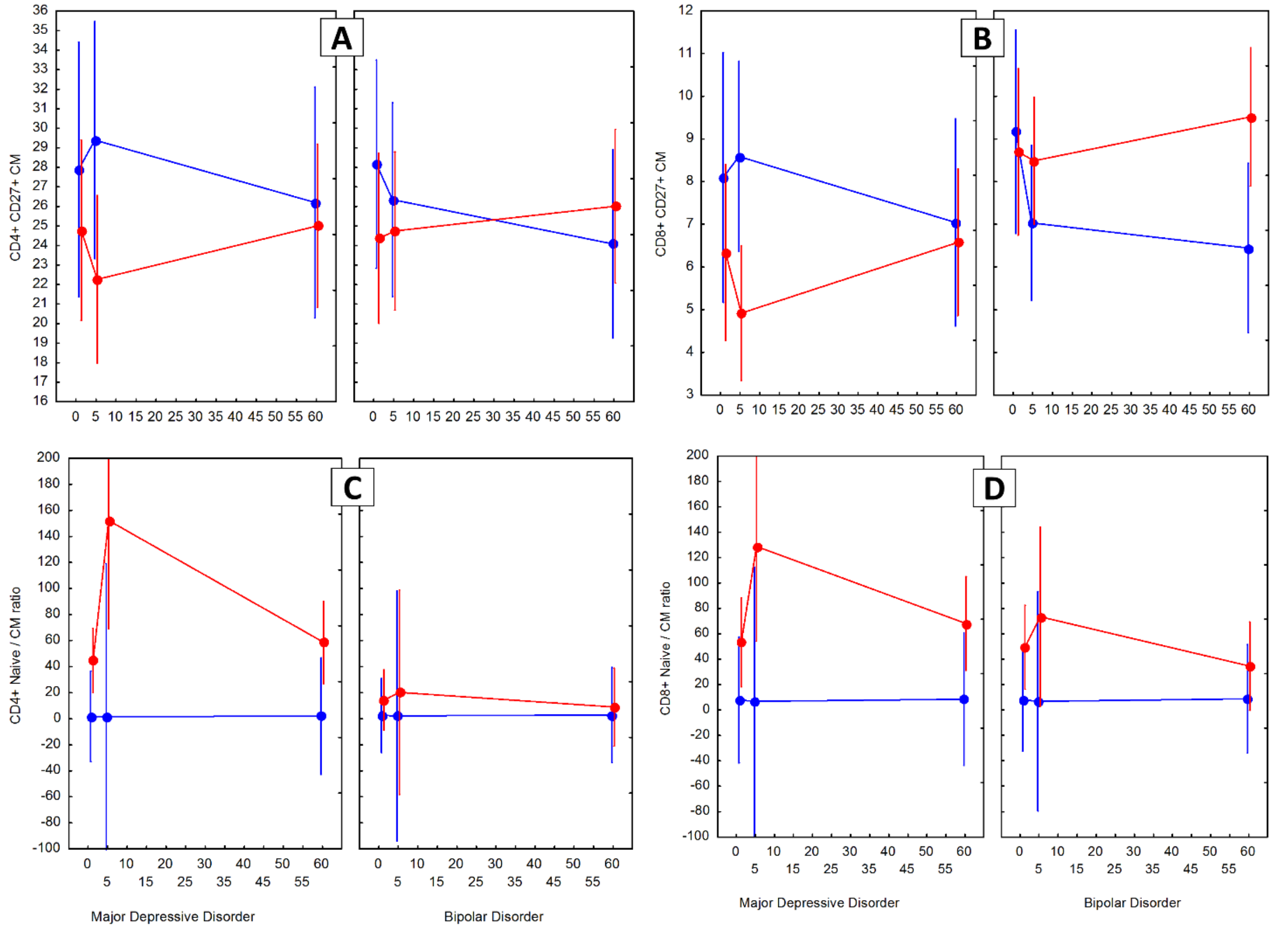

**Supplementary Figure 3**

Seum levels of sCD25, CRP, IL-7 and BDNF. Red=Aldesleukin; Blue=Placebo. Points are means, whiskers are SEM.

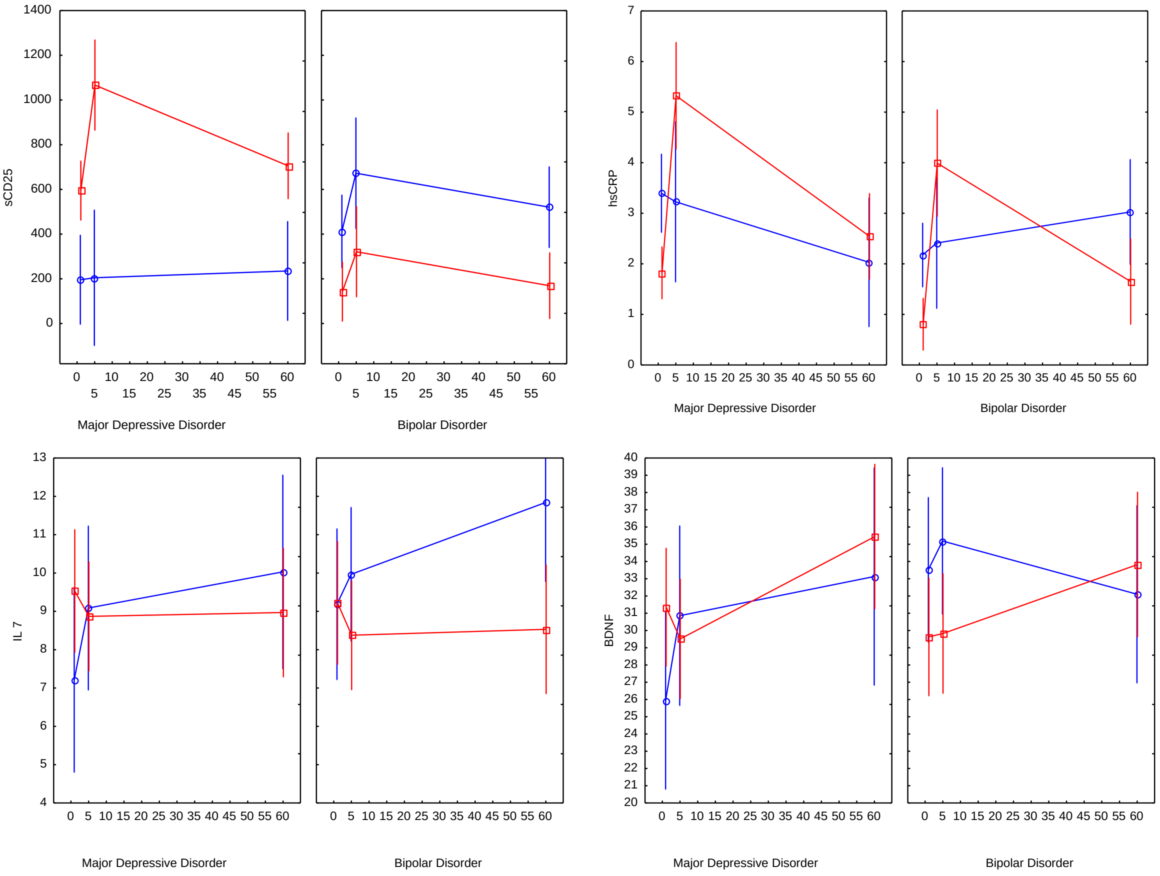
